# Durability of the Benefit of Vagus Nerve Stimulation in Markedly Treatment-Resistant Major Depression: A RECOVER Trial Report

**DOI:** 10.1101/2025.10.15.25337077

**Authors:** Charles R. Conway, A. John Rush, Scott T. Aaronson, Mark T. Bunker, Charles Gordon, Mark S. George, Patricio Riva-Posse, Rebecca M. Allen, Ziad Nahas, Christopher L. Kriedt, John Zajecka, David L. Dunner, João Quevedo, Yvette Sheline, Walter Duffy, Brian J. Mickey, Mary Stedman, Gustavo Alva, Lucian Manu, Quyen Tran, Charles F. Zorumski, Matthew Macaluso, Michael Banov, Cristina Cusin, Jeffrey I. Bennett, Hunter Brown, Jeffrey Way, Olivia Shy, Ying-Chieh (Lisa) Lee, R. Hamish McAllister-Williams, Roger S. McIntyre, Harold A. Sackeim

## Abstract

**Importance:** Greater levels of treatment resistance in major depressive disorder (MDD) are associated with lower rates of initial benefit and higher rates of relapse (lower durability).

**Objective:** Characterize depressive symptoms, function, and quality of life (QoL) over 24 months of adjunctive vagus nerve stimulation (VNS) in participants with markedly treatment-resistant depression.

**Design:** Prospective, open-label, single-arm, long-term extension study (RECOVER) conducted from September 2019 to April 2025.

**Setting:** Outpatient.

**Participants:** Adults with moderate-severe MDD with ≥4 failed antidepressant trials in the current episode, randomized to blinded, adjunctive VNS for 12 months, who subsequently received open-label, adjunctive VNS for 12 additional months (N=214).

**Interventions:** VNS and concomitant psychotropic medications and interventional psychiatric modalities (electroconvulsive therapy, transcranial magnetic stimulation, ketamine/esketamine) were characterized over the 12-month extension.

**Main Outcomes and Measures:** The durability of benefit achieved at 12 months was assessed at 18 and 24 months for depressive symptoms (3 scales), daily function, QoL, a tripartite composite of all 3 domains, and the Clinical Global Impression–Improvement (CGI-I) scale (overall improvement). Loss of benefit and relapse were assessed, along with the emergence of meaningful benefit in participants without benefit at 12 months. Substantial benefit (at least 50% symptom reduction from baseline; CGI-I of 1 or 2; tripartite with at least 2 of 3 subscales evidencing benefit) and meaningful benefit thresholds for symptoms (at least 30% reduction from baseline), function, QoL, CGI-I, and the tripartite measure were set *a priori*.

**Results:** Most participants with substantial benefit maintained their benefit (18-month median=78.8%; 24-month median=79.0% across 5 measures), as did participants with at least meaningful benefit at 12 months (18-month median=83.1%; 24-month median=81.3% across 7 measures). Furthermore, many participants with no meaningful benefit at 12 months achieved it at 18 (median=30.6%) and 24 (median=37.8%) months. The strong maintenance of benefit was not accounted for by changes in psychotropic medications or interventional psychiatry modalities.

**Conclusions and Relevance:** Depressive symptom, daily function, and QoL benefits obtained after 12 months of adjunctive VNS were sustained in about 80% of participants continuing VNS. Approximately 30% with no meaningful benefit at 12 months accrued increased benefit over the subsequent year.

**Significance Statement:** Patients with markedly treatment-resistant major depression have low likelihood of benefiting from antidepressant treatments and high likelihood of rapid relapse if benefit is obtained. In 214 participants randomized in the RECOVER trial to active treatment with adjunctive Vagus Nerve Stimulation (VNS) therapy, outcomes after a second year of stimulation were examined for depressive symptom severity, function, and quality of life. Participants with meaningful benefit at 12 months maintained this level of benefit or better at 24 months at high rates (median ≥80% across all measures). Durability was strong for symptoms, function, and quality of life regardless of the threshold used to define benefit and was not attributable to changes in concomitant treatments. Moreover, a substantial proportion of participants had meaningful improvement at 24 months who did not at 12 months. In a sample characterized by profound chronicity and treatment resistance, active VNS was associated with exceptionally strong durability of benefit.

## INTRODUCTION

Most patients with major depressive disorder (MDD) substantially improve with 1 to 3 antidepressant treatment attempts, but a sizable proportion (18%–33%) do not.^1^ Individuals with treatment-resistant depression (TRD)^2^ or difficult-to-treat depression^3^ have higher rates of relapse,^1^ hospitalization,^4^ suicide attempt,^5^ medical morbidity,^6^ and disability^7^ than those with more treatment-responsive depression.

TRD presents 2 fundamental therapeutic challenges: initially achieving benefit and subsequently sustaining it. The likelihood of achieving acute symptomatic improvement decreases with increasing levels of treatment resistance.^1,2,8,9^ Also, when an initial benefit is achieved, the likelihood that it is sustained decreases with higher levels of treatment resistance.^1,2,10^ This finding was prospectively documented in the STAR*D trial, where, following 2 inadequate treatment responses, the likelihood of achieving sustained remission for 1 year was less than 5% with subsequent medication regimens, due both to low rate of acute improvement and poor durability of benefit.^1,11^ As another example, electroconvulsive therapy (ECT) produces remission in 50% to 70% of patients with TRD,^12^ but approximately 50% of these patients relapse (most within 2 to 3 months).^13,14^ Even with maintenance ECT, less than half of patients with TRD retain remission status at 12 months.^13,15^ For TRD, and especially markedly TRD, an assessment of the durability of the initial benefit is critical when evaluating the clinical impact of treatment in patients where loss of initial benefit is highly likely.

Cervically implanted vagus nerve stimulation (VNS) has shown promise in achieving meaningful antidepressant effects^16,17^ and sustaining this benefit over time.^17,18^ Previous VNS trials found that approximately 40% to 50% of patients with markedly TRD show meaningful improvement 3 to 12 months after starting stimulation,^17,19^ a much longer interval to benefit than observed with other antidepressant treatments.^20^ Critically, early VNS studies documented that a large proportion of patients sustained substantial benefit for at least 1 to 2 years.^18,21^ Similarly, using registry data, Aaronson et al.^17^ found that individuals classified as responders to VNS adjunctive to treatment as usual (TAU) had a significantly longer time to relapse or recurrence compared with patients who met response criteria while treated with TAU alone. Determining whether the initial benefits of adjunctive VNS are indeed durable is essential.

In markedly TRD, in addition to durability, the breadth and nature of initial and sustained benefits requires consideration.^22^ Traditionally, therapeutic trials in MDD examine change in depression symptom severity as the sole outcome domain to characterize short-term benefit and its consequent durability.^23^ Patients with markedly TRD, however, have profound deficits in day-to-day function and quality of life (QoL), often disproportionate to symptom severity.^24–26^ The initial acquisition of benefits in these domains, as well as their durability, have rarely been described in such patients.^27^

Further, the threshold defining clinical benefit has traditionally corresponded to response, typically a ≥50% reduction relative to baseline, or remission, the virtual absence of symptoms.^28,29^ In markedly TRD, these outcomes are relatively rare and a lesser degree of improvement, corresponding to partial response (PR; e.g., ≥30% reduction), is associated with meaningful improvement in QoL.^30,31^ Partial response has also shown greater sensitivity to change over time and to treatment group differences compared with the traditional outcome categories.^32,33^ However, the clinical utility of adopting lower improvement thresholds for detecting clinical benefit would be questionable if the benefits were not sustained. No previous study has examined durability of benefit in patients with MDD using meaningful benefit (partial symptom response) and substantial benefit (symptom response) levels of improvement.

This report describes clinical outcomes after 18 and 24 months of stimulation for individuals who were randomized to blinded treatment with *active* VNS combined with TAU in the initial 12 months of the large multisite RECOVER trial (NCT03887715).^34^ Outcome domains included measures of depressive symptoms, daily function, and QoL. In addition, outcomes were examined using a tripartite composite measure that combines these 3 domains into a single metric.^35,36^ For all outcomes, thresholds were defined as denoting “meaningful benefit” (MB) (Table 1). Meaningful benefit corresponded to PR for depressive symptom measures and the *a priori*, previously empirically determined minimal clinically important differences (MCID) for daily function and QoL measures, to 1 or more domains with at least MB on the tripartite composite measure, and to a score of 3 or less on the Clinical Global Impression–Improvement (CGI-I) scale. A greater degree of improvement (i.e., substantial benefit [SB]) was defined using the traditional threshold for response on the depressive symptom measures, 2 or more domains with at least MB for the tripartite metric, and a CGI-I score of 1 or 2. Substantial benefit was not defined for the function and QoL measures, given the absence of investigation justifying a cutoff beyond the MCID. Durability of benefit was described separately for groups that manifested meaningful or substantial benefit at 12 months. This study compared clinical outcomes at 12, 18, and 24 months and determined whether clinical outcomes improved, deteriorated, or were unchanged during the second year of stimulation. Participants were classified in terms of their outcome status at 12 months, and durability was assessed by the percentage of participants continuing to manifest benefit at 18 or 24 months. Similarly, this study also documented the percentages of participants who lost benefit (relapse) or achieved benefit (late improvement) during the second year. This study addressed the following specific questions:

1) For each outcome measure, how did outcomes at 18 and 24 months compare with those at 12 months? What proportion of participants was classified as having no benefit, meaningful benefit, or substantial benefit at the completion of the second year relative to the proportions after the first year of stimulation?
2) Were the benefits obtained at 12 months durable? Did a substantial proportion of participants who benefited after 1 year of VNS sustain the benefit over the following year?
3) Did a substantial proportion of participants lose benefit over time? What proportion of participants who achieved meaningful or substantial benefit after 1 year of VNS lost that benefit during the following year?
4) For participants with no benefit after 1 year of VNS, what proportion developed meaningful or substantial benefit over the following year?
5) Are the findings regarding longer-term outcomes contingent on outcome domain, or are they consistent across depressive symptom, daily function, QoL, and tripartite measures?
6) Are the durability and relapse findings contingent on the degree of benefit at 12 months? Is meaningful benefit at 12 months informative of participant outcomes at 24 months, and is meaningful benefit durable?

This study described outcomes during the second year of treatment with active VNS and unconstrained TAU. To examine whether the durability findings could be attributed to change in concomitant treatment regimens, the number and classes of psychotropic medications each participant reported at 12 months and last follow-up (18 or 24 months) were compared, as was exposure to interventional psychiatry treatments (ECT, transcranial magnetic stimulation [TMS], and esketamine/ketamine). Hence, the study also addressed this question:

7) How did the number and type of concomitant medications and use of interventional psychiatry modalities change over the course of the second year? Could such changes account for findings regarding durability of benefit?

**Table 1.** Summary of the measures and thresholds used to define categories of benefit.

|  | **Meaningful Benefit (MB)** | **At Least Meaningful Benefit (≥MB)** | **Substantial Benefit (SB)** | **Remission** |
| --- | --- | --- | --- | --- |
| **MADRS** | 30%–49% reduction from baseline | ≥30% reduction from baseline | ≥50% reduction from baseline | Score ≤9 |
| **QIDS-C** | 30%–49% reduction from baseline | ≥30% reduction from baseline | ≥50% reduction from baseline | Score ≤5 |
| **QIDS-SR** | 30%–49% reduction from baseline | ≥30% reduction from baseline | ≥50% reduction from baseline | Score ≤5 |
| **CGI-I** | Score=3 | Score ≤3 | Score ≤2 | Score=1 |
| **Mini-Q-LES-Q** | ≥11.89% increase from baseline | NA | NA | NA |
| **WPAI item 6** | ≥2-point reduction from baseline | NA | NA | NA |
| **Tripartite metric<sup>a</sup>** | Score=1 | Score ≥1 | Score ≥2 | NA |

For symptom measures (MADRS, QIDS-C, QIDS-SR, and CGI-I), meaningful benefit is equivalent to partial response; substantial benefit is equivalent to response.
<sup>a</sup>Score is count of outcome domains that manifested meaningful benefit.
Abbreviations: CGI-I, Clinical Global Impression–Improvement; MADRS, Montgomery-Åsberg Depression Rating Scale; Mini-Q-LES-Q, 7-item subset of the Quality of Life Enjoyment and Satisfaction Questionnaire; NA, not applicable; QIDS-C, Quick Inventory of Depressive Symptomatology–Clinician; QIDS-SR, Quick Inventory of Depressive Symptomatology–Self-Report; WPAI, Work Productivity and Activity Impairment Questionnaire.

## METHODS

### Study Design

The first phase of the RECOVER trial (ClinicalTrials.gov Identifier: NCT03887715) used a multicenter, randomized, triple-blind, sham-controlled design to evaluate the safety and effectiveness of adjunctive VNS over a 12-month period in participants with markedly TRD.^34^ Participants received either adjunctive active or sham VNS therapy. TAU was continued throughout the study. All participants provided written informed consent, and the study was approved by the local Institutional Review Board of the 84 participating US sites. The study procedures were in accordance with the ethical standards of the Helsinki Declaration.

After the 12-month blinded, randomized phase, all participants were treated openly with active VNS and TAU, without constraint on changes in VNS parameters or concomitant treatments. This report describes the depressive symptom, daily function, and QoL outcomes during the second year of stimulation in the 214 participants randomized to the active VNS arm for the first 12 months and who completed outcome evaluations at 18 or 24 months. Participants initially randomized to the sham VNS arm were excluded, as they had limited exposure to active VNS, an intervention with long delay to peak effectiveness, and durability of the benefit achieved with a 12-month sham intervention has limited practical import.

### Patient Sample

Participants were ≥18 years of age; in a major depressive episode (MDE) of MDD based on *Diagnostic and Statistical Manual of Mental Disorders, Fifth Edition* (DSM-5) criteria^37^; had ≥4 failed, adequate antidepressant treatments in the current MDE^8^; and had at least moderately severe depression at baseline (≥22 on the Montgomery-Åsberg Depression Rating Scale [MADRS]^38^ on 2 occasions, 14–16 days apart). Baseline MADRS severity was the average of these 2 assessments.

A total of 249 participants were initially randomized to active VNS. Of these, the following were not included in the analytic sample for this report: 15 participants were implanted but discontinued prior to the first 3-month assessment; an additional 13 participants continued after the 3-month assessment but did not have the 12-month assessment used to classify trial outcomes; 7 participants missed the 18- and 24-month assessments. These exclusions resulted in a final analytic sample of 214, which represented 85.9% (214/249) of participants initially randomized to VNS plus TAU (VNS ON+TAU). This high retention rate suggests that differential dropout or selection bias had minimal impact on longer-term outcomes.

This is a naturalistic, observational study of longer-term outcomes of a cohort expected to have especially poor durability of benefit given the sample’s profound chronicity of illness and treatment resistance. The major analyses regarding change in outcome rates during the second year of stimulation were conducted in a subsample (N=185) that had outcome assessments at 12, 18, and 24 months without missing observations, allowing within-group statistical comparison. The findings from the larger primary sample (N=214), based on all available follow-up data, are reported in Table S1.

### Outcome Assessments

Depressive symptom severity was assessed by the clinician-rated MADRS and Quick Inventory of Depressive Symptomatology–Clinician (QIDS-C) at the clinic visits. Both were completed by trained, off-site, telephone-based raters masked to protocol and treatment. Participants completed the Quick Inventory of Depressive Symptomatology–Self-Report (QIDS-SR) at the same clinic visits. The MADRS assesses 10 symptoms, each rated 0 to 6 over the previous week.^38^ The QIDS-C and QIDS-SR assess 9 depressive symptom domains (each rated 0 to 3) that define an MDE based on DSM-5 criteria over the previous week.^39^ For each symptom scale, PR and response were defined as ≥30% or ≥50% reduction in the baseline total score, respectively, based on independent studies that established these thresholds as clinically meaningful.^35^ On these scales, the terms partial response and meaningful benefit (MB) are equivalent, as are response and substantial benefit (SB). Remission was defined as a score of ≤9 for the MADRS and ≤5 for both the QIDS-C and QIDS-SR. Table 1 defines meaningful benefit, substantial benefit, and remission for each scale.

General psychiatric status was assessed at baseline with the Clinical Global Impression–Severity (CGI-S) scale over the previous week based on behavior, symptoms, and function.^40^ The CGI-I scale, completed by on-site blinded psychiatrists and psychologists, assessed the participants’ overall clinical condition compared with the baseline CGI-S score.^40^ At each assessment, on the CGI-I, an MB was defined as a score of ≤3 (at least “minimally improved”), while an SB was defined as a score of ≤2 (at least “much improved”), and remission was defined as a score of 1, based on research validating these cutoffs.^41^

The Work Productivity and Activity Impairment Questionnaire (WPAI) measures absenteeism, presenteeism, and productivity for working participants (items 2–5).^42–44^ Since most participants (74.6%) were not employed,^45,46^ WPAI items 2 through 5 did not apply. WPAI item 6 assesses, over the previous week, the effect of depression (from 0 for “no effect” to 10 for “massive effect”) on regular daily activities (work around the house, shopping, childcare, exercising, studying, etc). An MCID or MB on item 6 was defined as a reduction of ≥2 points based on research validating this cutoff.^35^

The 7-item, self-report subset of the Quality of Life Enjoyment and Satisfaction Questionnaire (Mini-Q-LES-Q)^47^ assessed QoL over the previous week using the items on the 14-item Q-LES-Q most sensitive to change during treatment of MDD: work, household activities, social relationships, family relationships, leisure time activities, ability to function in daily life, and overall sense of well-being. Each item was rated from 1 (very poor) to 5 (very good) then converted to percent maximum scores by subtracting 7 from the raw total score (range=7–35) and dividing by 28. The MCID for the Mini-Q-LES-Q was estimated *a priori* as an 11.89% increase from baseline based on previous research using the 14-item Q-LES-Q.^48^ Participants meeting this threshold were classified as manifesting MB in QoL, while those below were classified as having no meaningful benefit (NB).

The tripartite composite combined measures of change in symptom severity (QIDS-C), function (WPAI item 6), and QoL (Mini-Q-LES-Q) into a single metric.^35^ Composite scoring ranged from 0 to 3, based on the number of domains in which at least meaningful benefit (≥MB) was manifest: scores of 0 were categorized as NB, 1 as MB, and 2 or 3 as SB. The QIDS-C was selected *a priori* as the depressive symptom component of the tripartite measure, as it was based on blinded, off-site clinician ratings and showed superior sensitivity to symptom change during the randomized phase compared with the MADRS.^33^ The tripartite metric was introduced in the RECOVER trial and has been shown to capture a wider group of patients than any one dimension alone. It correlates substantially with clinician global judgment of improvement and has shown utility in identifying the patient features predictive of VNS outcome.^35,36^ Durability of benefit has rarely been examined for daily function and QoL outcomes, and never for this tripartite metric.

All baseline assessments were completed before randomization.^34^ During the initial 12 months following randomization, depressive symptoms were assessed monthly, from months 3 through 12, and function and QoL were assessed quarterly. All 3 domains were assessed at 18 and 24 months during the second year of the trial.

### Statistical Analyses

This was an observational, naturalistic study of the durability and emergence/loss of benefit in the second year of the RECOVER participants initially randomized to active VNS. Analyses were conducted on 7 outcome measures with 3 follow-up time points (12, 18, and 24 months). Confidence intervals and *P*-values were not adjusted for multiplicity (α = 0.05). Statistical analyses were performed with SAS 9.4 (SAS Institute, Cary, NC).

The study reports descriptive information on second-year categorical outcomes for participants with MDD who were originally randomized to the VNS ON+TAU study arm. At each assessment (12, 18, and 24 months), outcomes for each depressive symptom scale (MADRS, QIDS-C, and QIDS-SR), the CGI-I, and the tripartite metric were classified as no meaningful benefit (NB), meaningful benefit (MB), substantial benefit (SB), and remission (with exception of remission for the tripartite metric). Daily function (WPAI item 6) and QoL (Mini-Q-LES-Q) measures were classified only dichotomously, as reflecting NB or MB.

#### Change in Outcomes Over Second Year

For each outcome measure, rates of NB, MB, SB, and remission, as well as at least MB (≥MB; MB or SB), were computed at the 12-, 18-, and 24-month assessments. Statistical significance of the difference between the rates at the 12- and 24-month time points was tested with McNemar’s exact test, with corresponding 95% CIs estimated by the Mantel-Haenszel method for dependent proportions, in the subsample with no missing data at all 3 assessments.

#### Durability of Benefit

For each outcome, durability was operationalized as the proportion of participants with ≥MB at the 12-month time point who were classified as ≥MB at the 18- and 24-month assessments. Studies of longer-term outcomes in MDD have typically restricted follow-up to patients meeting response or remission criteria at an acute end point, including previous durability studies of VNS.^17,18^ To match these thresholds, the durability analyses were repeated, restricting the sample to those who met SB or remission criteria at the 12-month time point, and documenting the percentages manifesting ≥MB, SB, and remission at follow-up.

#### Loss of Benefit and Relapse

For each measure, the primary analyses of durability reflected the proportion of participants who manifested MB or SB at 12 months who retained ≥MB at follow-up. The first assessment of loss of benefit examined the inverse, the proportion of participants with MB or SB at 12 months who had NB at follow-up. However, this analysis likely overestimated meaningful loss of benefit, as a minor change in scores could result in a shift from MB to NB. In contrast, relapse is traditionally defined as a marked increase in symptomatology in patients who previously substantially benefited from an intervention.^14,18,29,49^ Therefore, we also applied a stricter definition of loss of benefit to the depressive symptom, CGI-I, and tripartite outcomes by calculating the proportion of participants with SB at 12 months who had NB at follow-up, operationalizing the traditional classification of relapse.

#### Emergence of Benefit

The foregoing analyses focused on the longer-term outcomes of participants who were classified with MB or SB at the 12-month assessment. Longer-term outcomes were also examined for those with NB at 12 months.

#### Concomitant Treatments

All ongoing medication and device-based treatments were documented at month 12 and last follow-up at 18 or 24 months and categorized according to the WHO Anatomical Therapeutic Chemical (ATC) Classification System.^50^ At each assessment, the total number of psychotropic medications (ATC Level 1 Nervous System) reported by the participant and the number per class (antianxiety, anticonvulsant, antidepressant, antipsychotic, anxiolytic, hypnotic, stimulant, and other) were calculated. Supplemental Methods 1 details the methods used in classifying medications. Paired t-tests determined whether overall psychotropic use and use of specific classes changed in the second year. The number of psychotropic medications added or subtracted at last follow-up relative to month 12 was computed for each class and the means tested against 0 with one-sample t-tests. Compared to other outcome measures, the tripartite metric had both the highest rate of participants manifesting benefit at 12 months and the strongest durability values. Accordingly, the analyses on change in medication use in the total sample were repeated to characterize the pattern specifically in the largest subgroup that exhibited sustained benefit. Similarly, exposure to interventional psychiatry treatments was tested for change over the study periods in the complete sample and for changes that occurred in the subgroup with sustained benefit. Similar analyses were performed using the CGI-I, which was selected because this metric also had a high rate of participants benefiting at 12 months and a high rate of sustained benefit at 24 months, and were conducted by blinded, on-site primary investigators with experience in TRD assessment.

## RESULTS

### Patient Sample

Table 2 details demographic, clinical, and treatment history characteristics of the participants with follow-up data at 18 or 24 months. RECOVER participants had profoundly chronic and markedly resistant depressive illness, with a mean of 13.5 lifetime antidepressant treatment trials with insufficient benefit, including a majority who had insufficient benefit from neurostimulation treatments in the current episode (52.3% TMS; 38.3% ECT). With a mean age of 55.2 years, the sample spent the majority of their lives (29.0 years; 52.6% lifetime) in depressive illness and averaged 17.1 years of sustained depression on entry into the trial.

**Table 2.** Demographic, clinical, and treatment history characteristics of the sample treated with VNS and TAU for 2 years (N=214)

| Variable |  |
| --- | --- |
| **Demographics** |  |
| Age, yr, mean (SD) | 55.2 (14.3) |
| Sex, N (%) |  |
| Female | 146 (68.2) |
| Male | 68 (31.8) |
| Employed, N (%) | 59 (27.6) |
| **Baseline Clinical Presentation** |  |
| Single-episode vs recurrent MDD, N (%) |  |
| Single-episode MDD | 95 (44.4) |
| Recurrent MDD | 119 (55.6) |
| MADRS total score (0–60), mean (SD) | 34.1 (4.9) |
| CGI-S score (1–7), mean (SD) | 5.1 (0.7) |
| QIDS-C total score (0–27), mean (SD) | 17.2 (3.1) |
| QIDS-SR total score (0–27), mean (SD) | 17.4 (3.7) |
| **Function and Quality of Life Assessments** |  |
| WPAI item 6, mean (SD) | 7.2 (2.0) |
| Q-LES-Q total score (14–70), mean (SD) | 34.2 (7.3) |
| Mini-Q-LES-Q total score (7–35), mean (SD) | 15.0 (4.1) |
| **Mood Disorder History, mean (SD)** |  |
| Age at onset of first symptoms of depression, years | 20.9 (12.1) |
| Age at definitive diagnosis of any major mood disorder, years | 27.1 (12.6) |
| Duration of current MDE, years | 17.1 (15.3) |
| Duration of lifetime MDEs, years<sup>a</sup> | 29.0 (16.5) |
| Percentage of lifetime in MDE<sup>b</sup> | 52.6 (24.4) |
| Lifetime number of prior hospital admissions for mood disorder | 2.3 (4.3) |
| Lifetime history of suicide attempts, n (%) | 89 (41.6) |

| **Antidepressant Treatments, Lifetime** |  |
| --- | --- |
| Lifetime number of inadequate responses to antidepressant treatments, mean (SD)<sup>c</sup> | 13.5 (9.2) |
| Lifetime number of inadequate responses to antidepressant pharmacotherapies, mean (SD) | 11.5 (8.6) |
| Lifetime history of ECT, N (%) |  |
| Responded | 25 (11.7) |
| Did not respond | 76 (35.5) |
| **Antidepressant Treatments in Current MDE** |  |
| Any pharmacotherapy, N (%) | 214 (100) |
| Esketamine | 48 (22.4) |
| Any psychotherapy, N (%) | 162 (75.7) |
| Any neuromodulation, N (%) | 144 (67.3) |
| TMS | 112 (52.3) |
| ECT | 82 (38.3) |

<sup>a</sup>Duration of lifetime MDEs is the number of years of previous and current major depressive episodes. If both previous and current episodes were missing, then duration of lifetime episodes was considered as missing. If either of the 2 was not missing, then duration of lifetime episodes was the sum of the non-missing number. The maximum duration of estimated lifetime episodes in years was set as age at baseline. If the number of hospital admissions or of lifetime episodes was more than 10, the number was capped at 10.
<sup>b</sup>Percentage of lifetime in MDE was calculated as “duration of lifetime MDEs” divided by “age at baseline.”
<sup>c</sup>Including pharmacotherapies, ECT, TMS, and psychotherapies.
Abbreviations: CGI-S, Clinical Global Impression–Severity; ECT, electroconvulsive therapy; MADRS, Montgomery-Åsberg Depression Rating Scale; MDD, major depressive disorder; MDE, major depressive episode; Mini-Q-LES-Q, 7-item subset of the Quality of Life Enjoyment and Satisfaction Questionnaire; QIDS-C, Quick Inventory of Depressive Symptomatology–Clinician; QIDS-SR, Quick Inventory of Depressive Symptomatology–Self-Report; Q-LES-Q, Quality of Life Enjoyment and Satisfaction Questionnaire; TAU, treatment as usual; TMS, transcranial magnetic stimulation; VNS, vagus nerve stimulation; WPAI, Work Productivity and Activity Impairment Questionnaire.

Additionally, the sample was severely negatively impacted by depressive illness. A total of 72.4% were unemployed, and participants self-reported very poor QoL (mean percent max Q-LES-Q Short Form score of 48.9%, which falls below the “severe impairment” line [50%] of the scale; a normal control/community normative score typically resides in the normal range of 75%–91% max score, with severe impairment being 2 SDs below the norm)^51,52^ and were functionally impaired (WPAI item 6 mean score of 7.2, which ranks more impaired than studies of non-resistant depression [4.76],^53^ chronic migraine [5.52],^54^ and rheumatoid arthritis [3.33]).^55^

### Overall Patient Course

Table 1 details the definitions of meaningful benefit (MB), at least meaningful benefit (≥MB), substantial benefit (SB), and remission for all applicable metrics. Table 3 presents the proportion of participants classified as having NB, MB, SB, and remission at 12, 18, and 24 months for the depressive symptom measures and CGI-I; the proportion of participants having NB, MB, and SB for tripartite outcomes; and those with NB and MB in function (WPAI item 6) and QoL (Mini-Q-LES-Q) outcomes for participants with assessments at all 3 time points (12, 18, and 24 months; N=181). At 12 months, there were considerable differences among the outcome measures in classification of MB or at least MB (≥MB), with 43.6% achieving ≥MB on the MADRS and 80.0% on the tripartite composite measure. The increase from months 12 to 24 in the percentage achieving ≥MB was significant for the MADRS and CGI-I measures, and marginally so for the QIDS-C. The specific increase in SB status was significant for the QIDS-C and CGI-I, and marginally so for the MADRS. Notably, the increase from months 12 to 24 in the proportion of participants achieving remission was significant for the MADRS. Table 3 shows that for all outcome measures other than the QIDS-SR, the proportion of participants with NB decreased over the course of the second year, while the rates of participants achieving ≥MB and SB increased. Table S1 presents the rates at the 12-, 18-, and 24-month assessments across the sample.

**Table 3.**
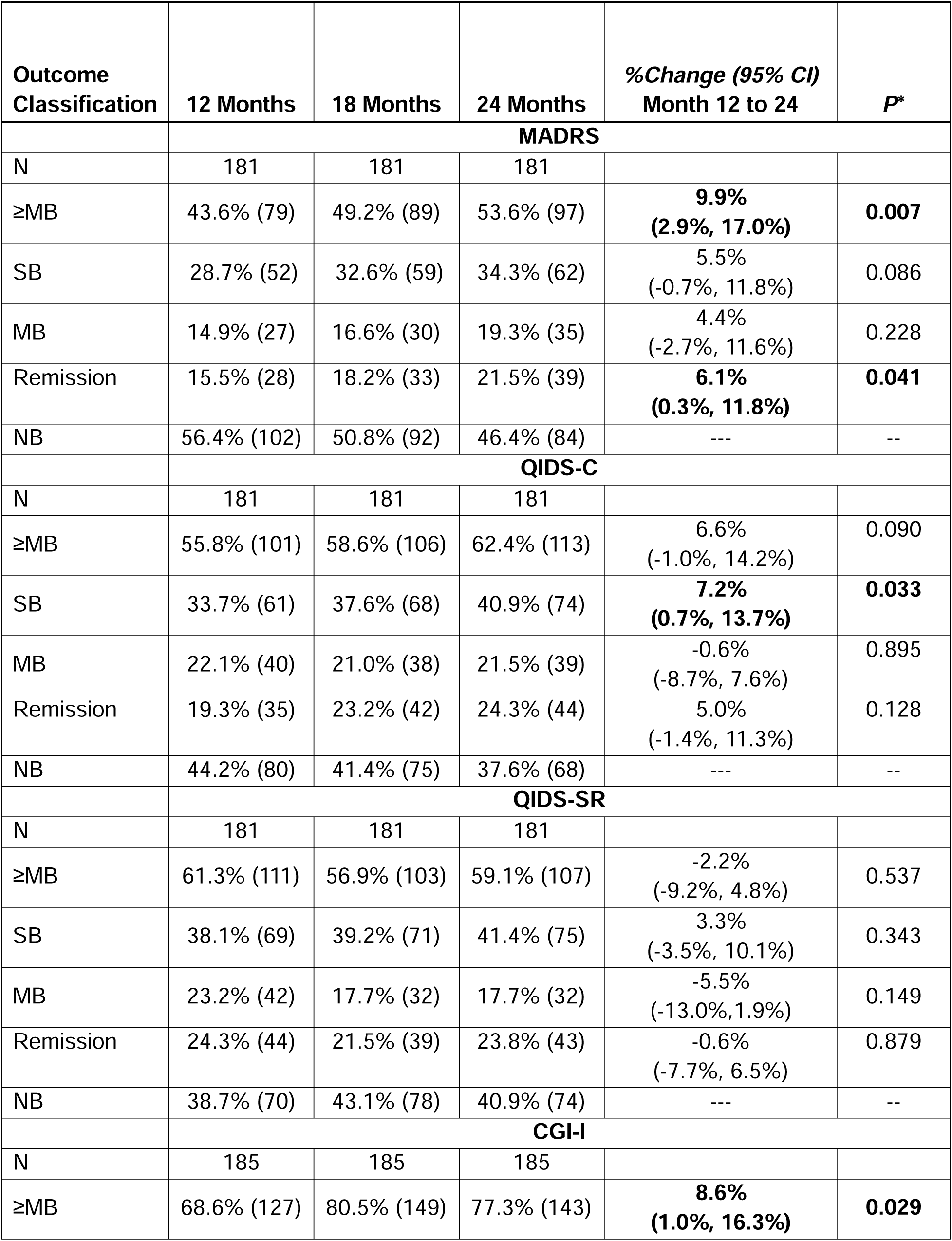
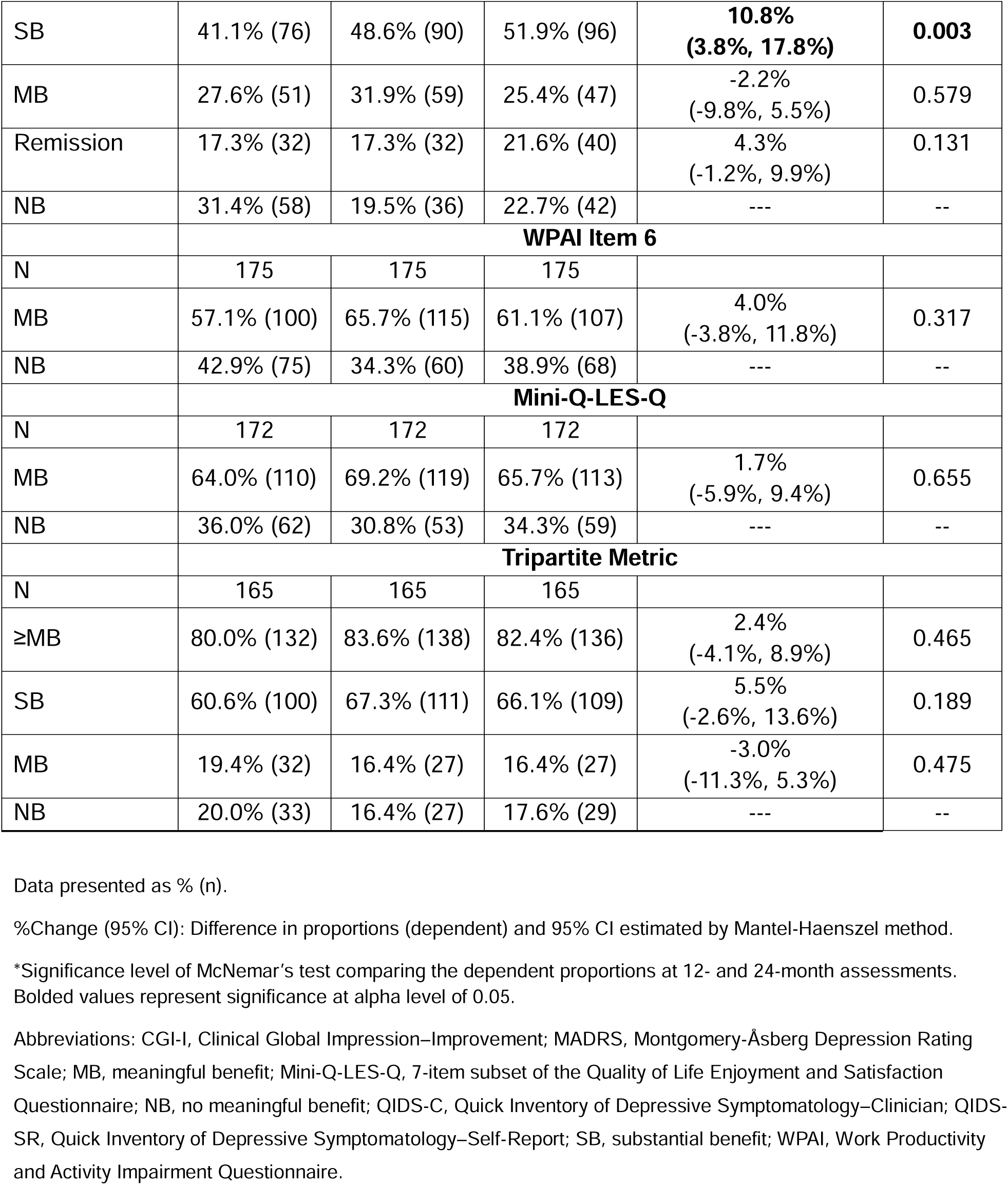
Rates of each clinical outcome at 12, 18, and 24 months in participants with no missing values at all 3 time points.

Missing data had minimal impact, as most participants remained in follow-up through 24 months. Discontinuation rates were similar regardless of month 12 outcomes, with a median dropout of 15.5% versus 17.7% for those with and without meaningful benefit, respectively (Table S2).

### Durability of Benefit

Table 4A and Figure 1 provide the percentage of participants manifesting MB or SB (≥MB) at the 12-month assessment who also manifested MB or SB at the 18- and 24-month assessments. At the 18-month assessment, these percentages ranged from 78.0% to 89.2% across all outcome measures. At 24 months, these percentages ranged from 79.2% to 89.6%. The median durability percentage across the 7 measures was 83.1% at 18 months and 81.3% at 24 months. Thus, across all outcomes, there was substantial retention of benefit during the second year of stimulation and little loss of durability from 18 to 24 months. A conservative analysis of all participants that imputed all missing data points as representing lost benefit found similar substantial retention of benefits (Table S3).

**Figure 1.**
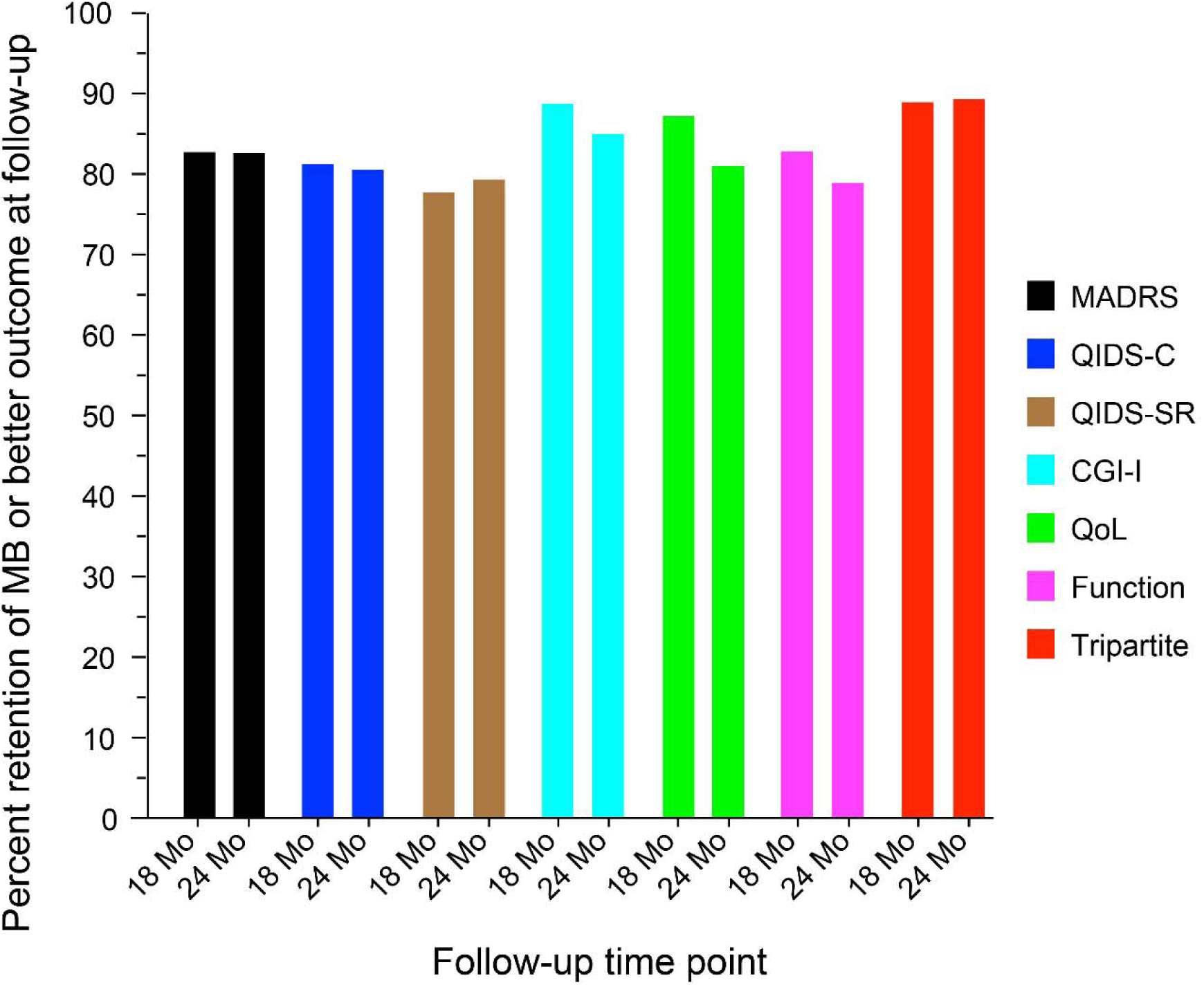
For each outcome measure, the percentage of participants who had at least meaningful benefit (≥MB) at 18 and 24 months of those who had at least meaningful benefit (≥MB) at 12 months Abbreviations: CGI-I, Clinical Global Impression–Improvement; MADRS, Montgomery-Åsberg Depression Rating Scale; MB, meaningful benefit; QIDS-C, Quick Inventory of Depressive Symptomatology–Clinician; QIDS-SR, Quick Inventory of Depressive Symptomatology–Self-Report; QoL, quality of life.

**Table 4.**
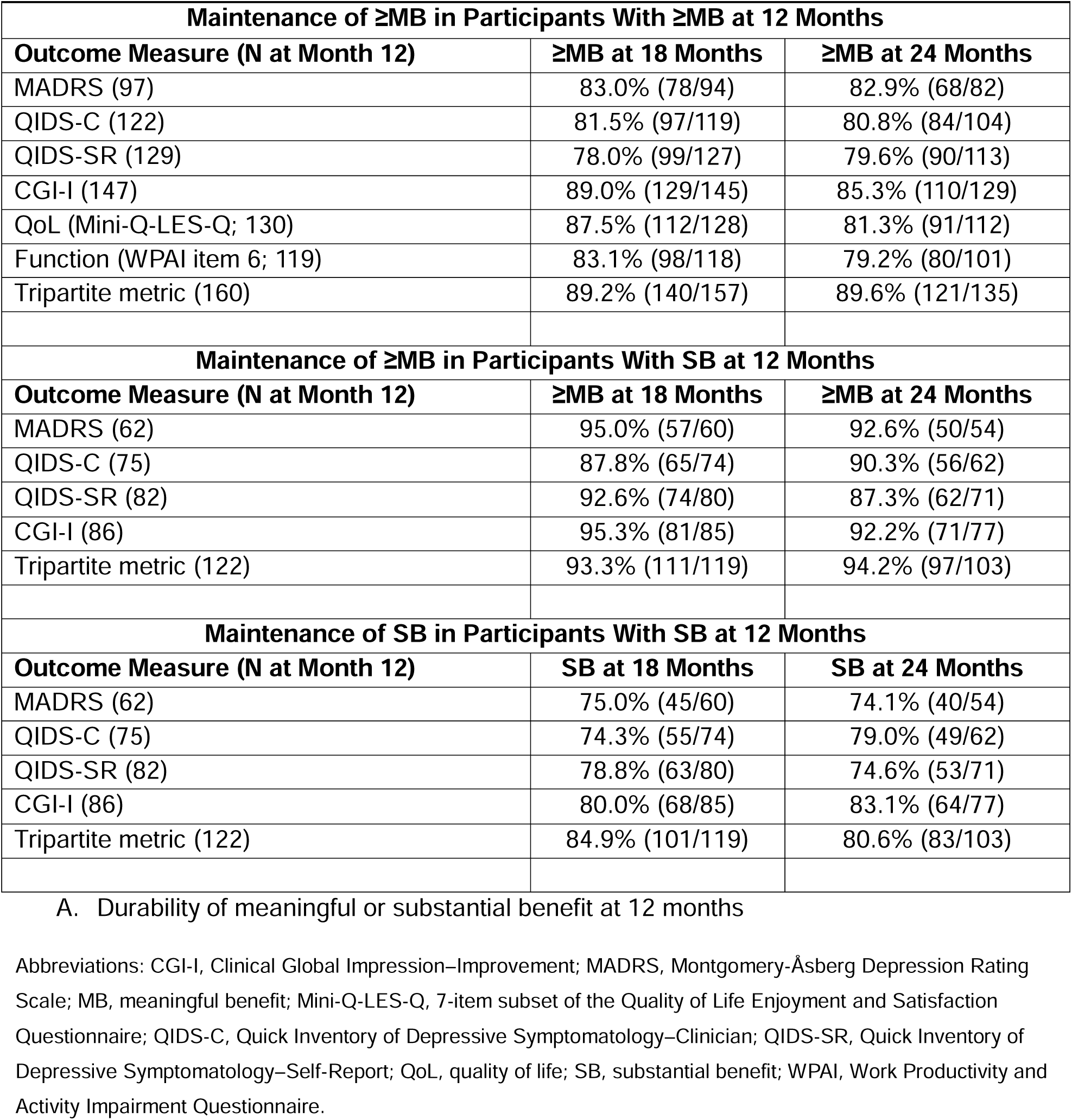
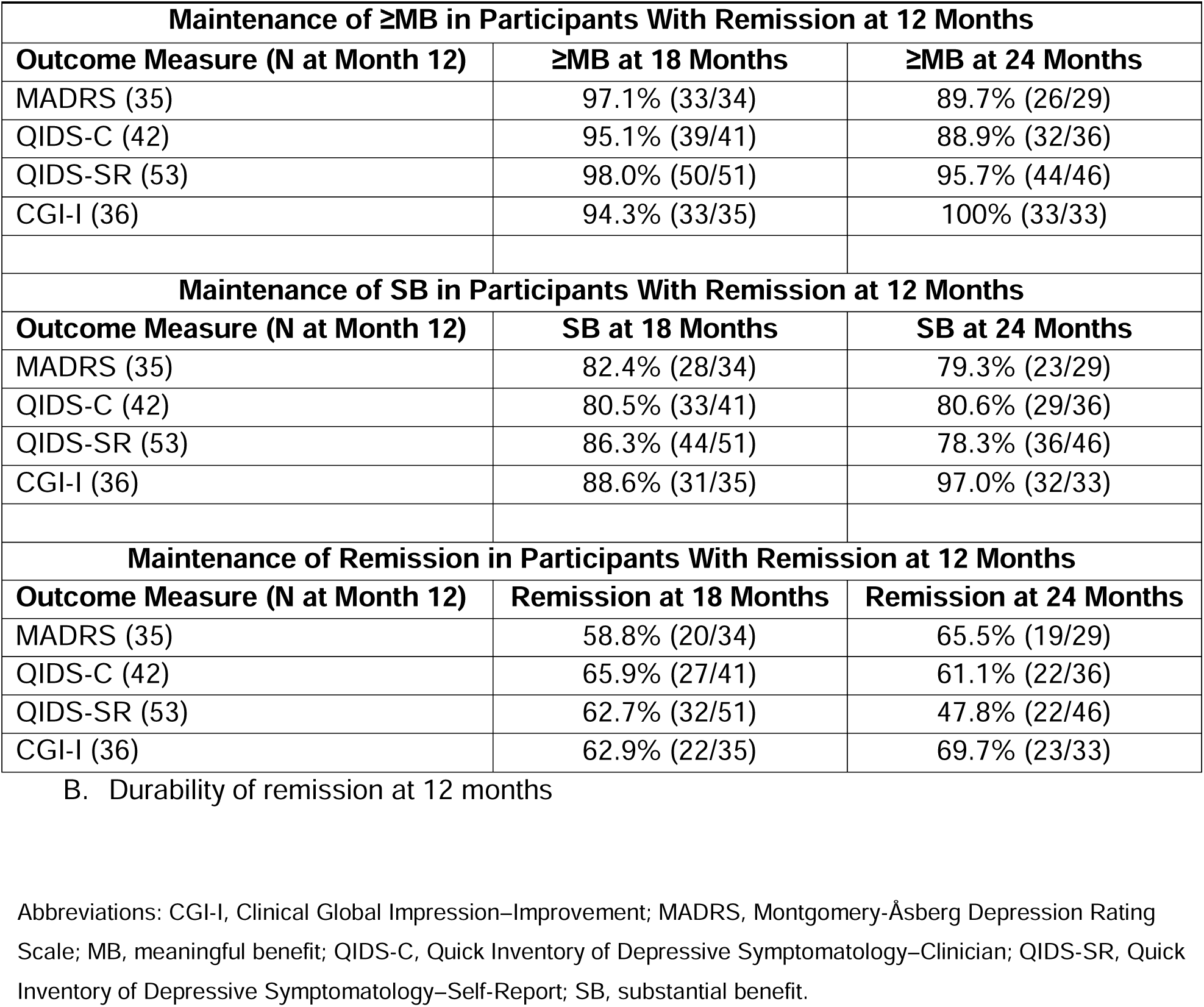
Percentage of participants who maintained (A) meaningful benefit or substantial benefit and (B) remission at 18- and 24-month follow-up.

| **Maintenance of ≥MB in Participants With ≥MB at 12 Months** |  |  |
| --- | --- | --- |
| **Outcome Measure (N at Month 12)** | **≥MB at 18 Months** | **≥MB at 24 Months** |
| MADRS (97) | 83.0% (78/94) | 82.9% (68/82) |
| QIDS-C (122) | 81.5% (97/119) | 80.8% (84/104) |
| QIDS-SR (129) | 78.0% (99/127) | 79.6% (90/113) |
| CGI-I (147) | 89.0% (129/145) | 85.3% (110/129) |
| QoL (Mini-Q-LES-Q; 130) | 87.5% (112/128) | 81.3% (91/112) |
| Function (WPAI item 6; 119) | 83.1% (98/118) | 79.2% (80/101) |
| Tripartite metric (160) | 89.2% (140/157) | 89.6% (121/135) |
| **Maintenance of ≥MB in Participants With SB at 12 Months** |  |  |
| **Outcome Measure (N at Month 12)** | **≥MB at 18 Months** | **≥MB at 24 Months** |
| MADRS (62) | 95.0% (57/60) | 92.6% (50/54) |
| QIDS-C (75) | 87.8% (65/74) | 90.3% (56/62) |
| QIDS-SR (82) | 92.6% (74/80) | 87.3% (62/71) |
| CGI-I (86) | 95.3% (81/85) | 92.2% (71/77) |
| Tripartite metric (122) | 93.3% (111/119) | 94.2% (97/103) |
| **Maintenance of SB in Participants With SB at 12 Months** |  |  |
| **Outcome Measure (N at Month 12)** | **SB at 18 Months** | **SB at 24 Months** |
| MADRS (62) | 75.0% (45/60) | 74.1% (40/54) |
| QIDS-C (75) | 74.3% (55/74) | 79.0% (49/62) |
| QIDS-SR (82) | 78.8% (63/80) | 74.6% (53/71) |
| CGI-I (86) | 80.0% (68/85) | 83.1% (64/77) |
| Tripartite metric (122) | 84.9% (101/119) | 80.6% (83/103) |

| **Maintenance of ≥MB in Participants With Remission at 12 Months** |  |  |
| --- | --- | --- |
| **Outcome Measure (N at Month 12)** | **≥MB at 18 Months** | **≥MB at 24 Months** |
| MADRS (35) | 97.1% (33/34) | 89.7% (26/29) |
| QIDS-C (42) | 95.1% (39/41) | 88.9% (32/36) |
| QIDS-SR (53) | 98.0% (50/51) | 95.7% (44/46) |
| CGI-I (36) | 94.3% (33/35) | 100% (33/33) |
| **Maintenance of SB in Participants With Remission at 12 Months** |  |  |
| **Outcome Measure (N at Month 12)** | **SB at 18 Months** | **SB at 24 Months** |
| MADRS (35) | 82.4% (28/34) | 79.3% (23/29) |
| QIDS-C (42) | 80.5% (33/41) | 80.6% (29/36) |
| QIDS-SR (53) | 86.3% (44/51) | 78.3% (36/46) |
| CGI-I (36) | 88.6% (31/35) | 97.0% (32/33) |
| **Maintenance of Remission in Participants With Remission at 12 Months** |  |  |
| **Outcome Measure (N at Month 12)** | **Remission at 18 Months** | **Remission at 24 Months** |
| MADRS (35) | 58.8% (20/34) | 65.5% (19/29) |
| QIDS-C (42) | 65.9% (27/41) | 61.1% (22/36) |
| QIDS-SR (53) | 62.7% (32/51) | 47.8% (22/46) |
| CGI-I (36) | 62.9% (22/35) | 69.7% (23/33) |

B. Durability of remission at 12 months
Abbreviations: CGI-I, Clinical Global Impression–Improvement; MADRS, Montgomery–Åsberg Depression Rating Scale; MB, meaningful benefit; QIDS-C, Quick Inventory of Depressive Symptomatology–Clinician; QIDS-SR, Quick Inventory of Depressive Symptomatology–Self-Report; SB, substantial benefit.

To determine whether durability was impacted by the extent of clinical benefit at 12 months, the follow-up data were reexamined, restricting the sample to participants with SB status at 12 months (i,e., ≥50% reduction from baseline in symptom measures; Table 4A). The percentage of participants who manifested MB or SB at 18 and 24 months was higher than when a lower threshold of improvement (MB) was used to define benefit at 12 months. Across all outcomes, for participants who manifested SB at 12 months, the durability percentages ranged from 87.8% to 95.3% at 18 months (median=93.3%) and from 87.3% to 94.2% at 24 months (median=92.2%). Thus, nearly all these participants retained ≥MB through 24 months. As also seen in Table 4A, durability of benefit remained high even when requiring that SB status be maintained to qualify as durable, with medians of 78.8% and 79.0% at 18 and 24 months, respectively. Therefore, attaining MB and/or SB at 12 months was associated with high likelihood of maintaining benefit over the second year of stimulation.

Table 4B describes the percentage of participants manifesting ≥MB, SB, or remission at 18 and 24 months who achieved remission at 12 months. Across all outcome measures, among participants who achieved remission at 12 months, 58.8% to 65.9% maintained that level of benefit at 18 months (median=62.8%) and 47.8% to 69.7% at 24 months (median=63.3%). Few participants achieving remission at 12 months lost all benefit, with retention of SB having medians of 84.4% and 80.0% at months 18 and 24, respectively, and retention of ≥MB in nearly the entire sample, with medians of 96.1% and 92.7%.

### Loss of Benefit and Relapse

Loss of benefit was defined as NB status at follow-up among participants who had achieved MB or SB (≥MB) status at 12 months (Table 5). At 18 months, this loss of ≥MB was observed across outcomes in 10.8% to 22.0% of participants, with a median of 16.9%. At 24 months, this loss of benefit characterized 10.4% to 20.8% of participants, with a median of 18.8%. When the extent of clinical benefit at 12 months was increased to SB, reflecting traditional definitions of clinical response and relapse,^28,29^ rates of loss of benefit (relapse) were consistently lower at both follow-up time points, with medians of 6.7% and 7.8% at 18 and 24 months, respectively. Similar findings in loss of benefit were observed among participants who achieved remission at 12 months, with medians of 3.9% and 7.3% at 18 and 24 months, respectively.

**Table 5.** Percentage of participants with loss of benefit at 18- and 24-month follow-up.

| **Loss of Benefit in Participants With ≥MB at 12 Months** |  |  |
| --- | --- | --- |
| **Outcome Measure (N at Month 12)** | **NB at 18 Months** | **NB at 24 Months** |
| MADRS (97) | 17.0% (16/94) | 17.1% (14/82) |
| QIDS-C (122) | 18.5% (22/119) | 19.2% (20/104) |
| QIDS-SR (129) | 22.0% (28/127) | 20.4% (23/113) |
| CGI-I (147) | 11.0% (16/145) | 14.7% (19/129) |
| QoL (Mini-Q-LES-Q; 130) | 16.9% (20/128) | 20.8% (21/112) |
| Function (WPAI item 6; 119) | 12.5% (16/118) | 18.8% (21/101) |
| Tripartite metric (160) | 10.8% (17/157) | 10.4% (14/135) |
| **Loss of Benefit in Participants With SB at 12 Months** |  |  |
| **Outcome Measure (N at Month 12)** | **NB at 18 Months** | **NB at 24 Months** |
| MADRS (62) | 5.0% (3/60) | 7.4% (4/54) |
| QIDS-C (75) | 12.2% (9/74) | 9.7% (6/62) |
| QIDS-SR (82) | 7.5% (6/80) | 12.7% (9/71) |
| CGI-I (86) | 4.7% (4/85) | 7.8% (6/77) |
| Tripartite metric (122) | 6.7% (8/119) | 5.8% (6/103) |
| **Loss of Benefit in Participants With Remission at 12 Months** |  |  |
| **Outcome Measure (N at Month 12)** | **NB at 18 Months** | **NB at 24 Months** |
| MADRS (35) | 2.9% (1/34) | 10.3% (3/29) |
| QIDS-C (42) | 4.9% (2/41) | 11.1% (4/36) |
| QIDS-SR (53) | 2.0% (1/51) | 4.3% (2/46) |
| CGI-I (36) | 5.7% (2/35) | 0% (0/33) |

Data presented as % (n).
Observed: Proportions were calculated using all available data at each visit.
Abbreviations: CGI-I, Clinical Global Impression–Improvement; MADRS, Montgomery–Åsberg Depression Rating Scale; MB, meaningful benefit; Mini-Q-LES-Q, 7-item subset of the Quality of Life Enjoyment and Satisfaction Questionnaire; NB, no meaningful benefit; QIDS-C, Quick Inventory of Depressive Symptomatology–Clinician; QIDS-SR, Quick Inventory of Depressive Symptomatology–Self-Report; QoL, quality of life; SB, substantial benefit; WPAI, Work Productivity and Activity Impairment Questionnaire.

### Emergence of Benefit for Those Without Benefit at 12 Months

Table 6 describes the longer-term outcomes of participants who manifested NB at 12 months. At both 18 and 24 months, and for each outcome measure, a substantial number of participants manifested clinically meaningful benefit (≥MB). At 18 months, this characterized 23.7% to 54.7% of participants across outcome measures, with a median of 30.6%. At 24 months, this improvement ranged from 26.0% to 60.7% across outcome measures, with a median of 37.8%.

**Table 6.** Percentage of participants with no meaningful benefit at 12 months who had no meaningful benefit, meaningful benefit, substantial benefit, and meaningful benefit or substantial benefit at 18- and 24-month follow-up.

| Outcome Measure | 18 Months |  |  |  | 24 Months |  |  |  |
| --- | --- | --- | --- | --- | --- | --- | --- | --- |
|  | NB | MB | SB | ≥MB | NB | MB | SB | ≥MB |
| MADRS, N=116 | 76.3% | 16.7% | 7.0% | 23.7% | 70.2% | 16.3% | 13.5% | 29.8% |
| QIDS-C, N=91 | 73.0% | 20.2% | 6.7% | 27.0% | 62.2% | 25.6% | 12.2% | 37.8% |
| QIDS-SR, N=84 | 75.3% | 21.0% | 3.7% | 24.7% | 74.0% | 15.1% | 11.0% | 26.0% |
| CGI-I, N=67 | 45.3% | 39.1% | 15.6% | 54.7% | 39.3% | 29.5% | 31.1% | 60.7% |
| QoL (Mini-Q-LES-Q), N=76 | 69.4% | 30.6% | – | 30.6% | 59.1% | 40.9% | – | 40.9% |
| Function (WPAI item 6), N=89 | 60.0% | 40.0% | – | 40.0% | 64.6% | 35.4% | – | 35.4% |
| Tripartite metric, N=40 | 48.6% | 24.3% | 27.0% | 51.4% | 47.2% | 25.0% | 27.8% | 52.8% |

Abbreviations: CGI-I, Clinical Global Impression–Improvement; MADRS, Montgomery-Åsberg Depression Rating Scale; MB, meaningful benefit; Mini-Q-LES-Q, 7-item subset of the Quality of Life Enjoyment and Satisfaction Questionnaire; NB, no meaningful benefit; QIDS-C, Quick Inventory of Depressive Symptomatology–Clinician; QIDS-SR, Quick Inventory of Depressive Symptomatology–Self-Report; QoL, quality of life; SB, substantial benefit; WPAI, Work Productivity and Activity Impairment Questionnaire.

### Stability of Meaningful Benefit Outcomes

Figure 2 provides an alluvial diagram graphically conveying the changes in outcomes over time for the CGI-I and tripartite metric. Figures S1 and S2 provide the alluvial diagrams for the remaining outcome measures. Of the 5 outcome measures that provided MB and SB outcomes at 24 months, 28.6% to 40.5% (median=35.7%) of participants classified as MB at 12 months improved to SB at 24 months, 25.0% to 35.7% (median=33.3%) lost benefit (NB), and 26.2% to 46.2% (median=31.0%) maintained MB a year later. Thus, approximately twice as many participants who achieved MB status at 12 months maintained or improved benefit at 24 months than lost benefit.

**Figure 2.**
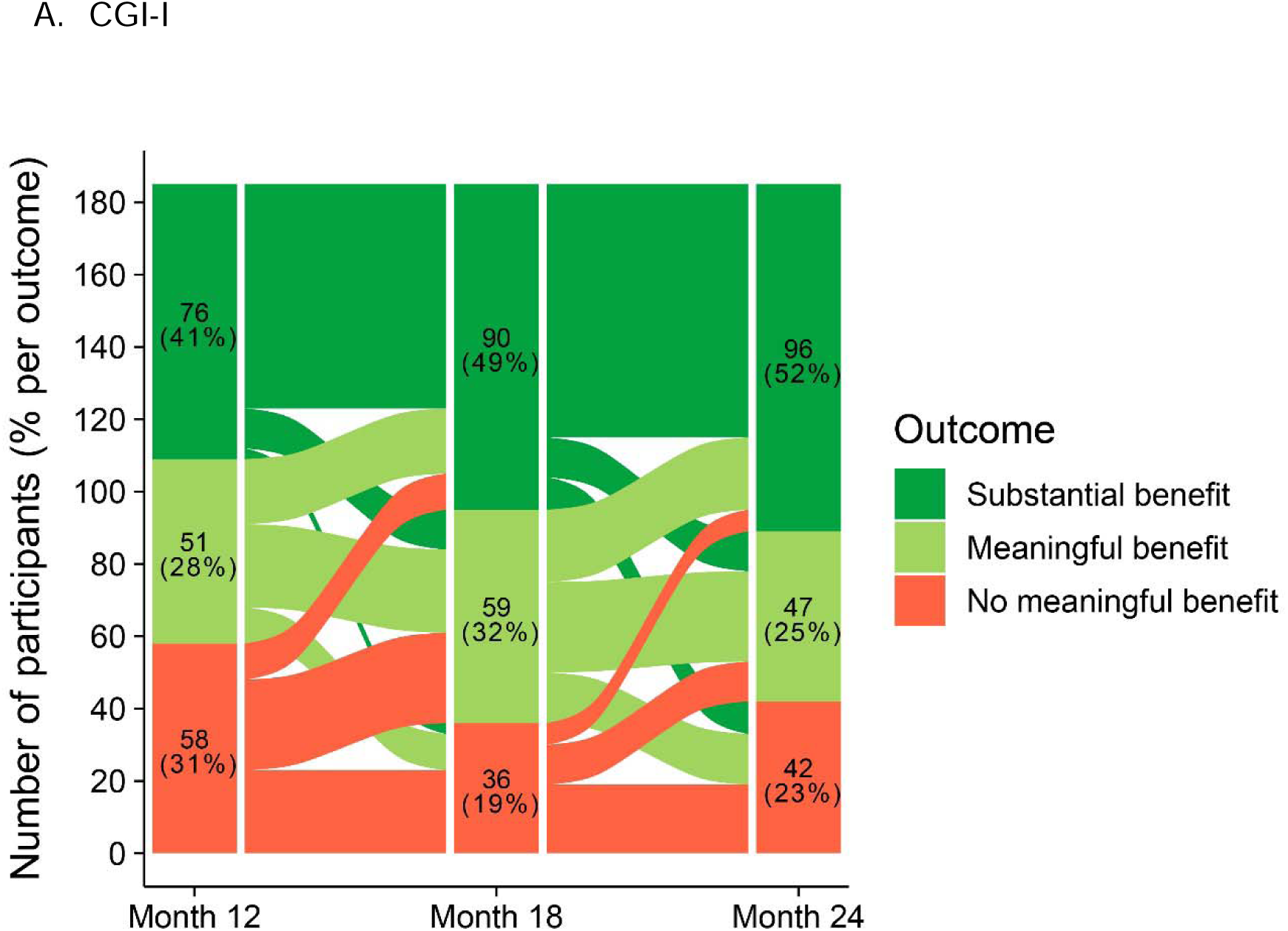
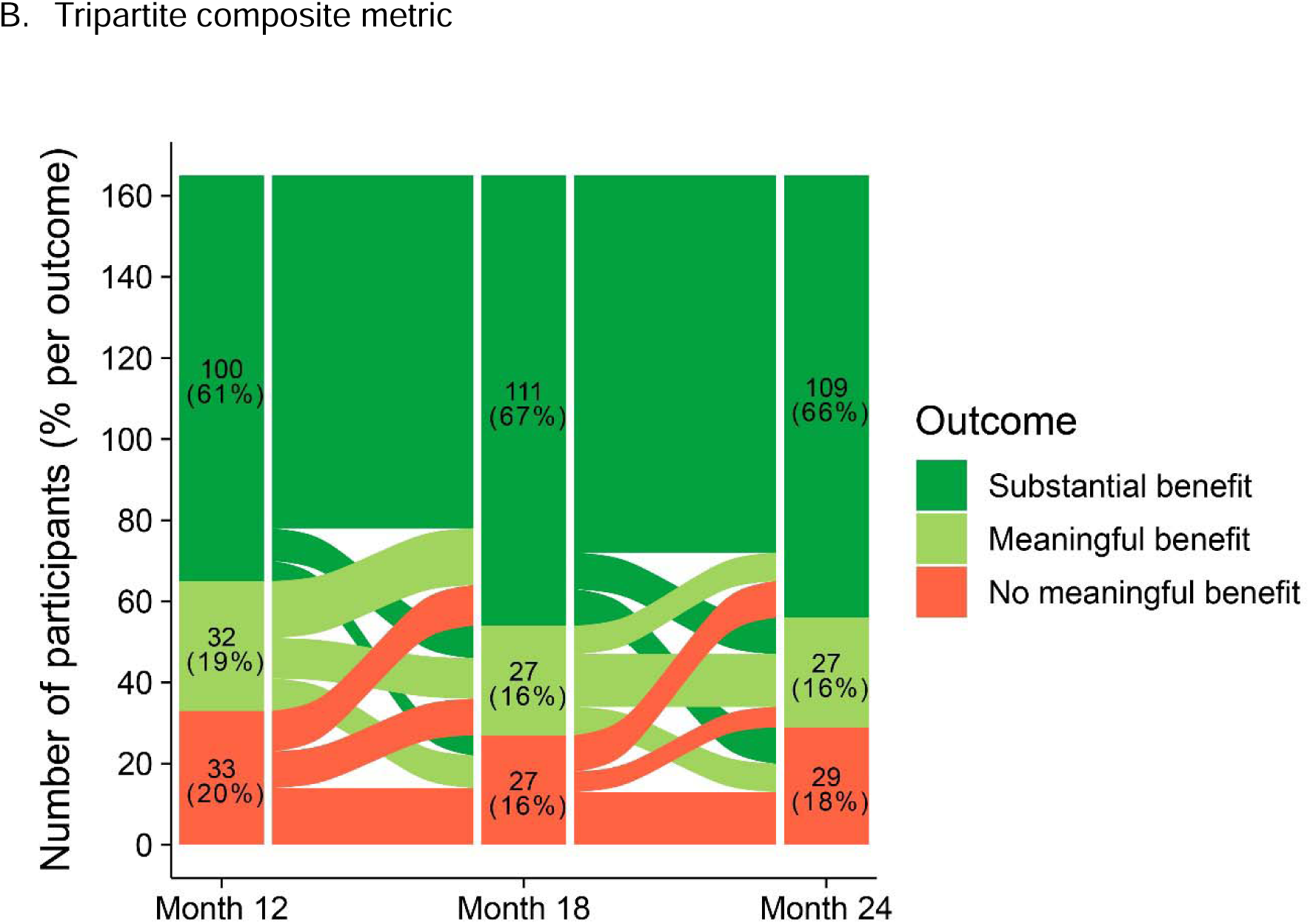
Alluvial diagram for changes in (A) CGI-I and (B) tripartite composite metric outcomes across months 12, 18, and 24 Abbreviation: CGI-I, Clinical Global Impression–Improvement.

### Concomitant Treatments

Table S4 summarizes the comparisons of the number and type of concomitant psychotropic medications reported at the 12-month and last follow-up (18 or 24 months) assessments. Across and within medication classes, paired t-tests were used to identify changes in number of concomitant psychotropics. These analyses were conducted in 3 samples: total sample (N=214), participants with durable MB on the tripartite metric (N=142; maintaining ≥1 on the tripartite), and participants with durable SB on the CGI-I, a more conservative metric (N=70; maintaining score of 1 or 2). None of the 24 comparisons (8 classes x 3 samples) reported in Table S4 achieved statistical significance. There was no evidence of consistent change in number of psychotropics administered over the follow-up period, either across or within classes. The mean number of psychotropics was 4.6 at month 12, and 4.6 at the last follow-up time point. This pattern held specifically in the subgroups that manifested durable benefit, regardless of definition.

Exposure to interventional treatments (ECT, TMS, and esketamine/ketamine) over the follow-up period was also examined in the 3 samples (Table S5). Overall, use of these interventions was quite limited, particularly ECT and TMS. For example, for the tripartite-durable subsample, while several participants continued maintenance ECT, TMS, or esketamine, no participants started a new course of ECT or TMS during the follow-up period, and 2 started a new course of esketamine/ketamine. Similar results were obtained in the CGI-I–durable subsample, as no participants received a new course of ECT or TMS, and 1 participant received a new course of esketamine/ketamine. Hence, the addition of interventional treatments was rare in the subgroups that manifested durable benefit, regardless of definition.

## DISCUSSION

The study participants had marked chronicity of depressive illness and profound treatment resistance with impairments of daily function and QoL. These clinical features have been consistently linked to poor prognosis in MDD.^2,56,57^ Therefore, achieving and sustaining meaningful clinical improvement would be considered unlikely in these patients. Nonetheless, this study documented strong durability of meaningful benefit in this markedly TRD sample treated with adjunctive VNS for 2 years. Across depressive symptoms, function, QoL, and the tripartite metric, and across off-site and on-site clinician and self-ratings, approximately 80% of participants with ≥MB at 12 months maintained this status at 18 and 24 months (Figure 1). Those participants with greater benefit at 12 months (substantial benefit [SB] or remission) maintained at least meaningful benefit (≥MB) at higher rates at each follow-up (Table 4A,B) compared to those with less robust benefit at 12 months. The rates of loss of benefit were notably low and mirrored the durability findings. Relapse was rare, as participants with more extensive improvement (SB or remission) at 12 months were especially unlikely to exhibit no meaningful benefit (NB) at each follow-up (Table 5). Across outcome measures, participants who had meaningful benefit (MB) at 12 months were twice as likely to maintain this level of benefit or improve to SB than manifest NB at the 24-month follow-up (Figure 2, Figures S1–S2). This finding underscores the utility of including partial response when classifying outcomes in markedly treatment-resistant MDD.^3,35^ Thus, strong durability was observed regardless of the threshold used to define benefit. Finally, though remission is a relatively rare outcome in markedly TRD, the data indicated that a meaningful subset (21.5%) of participants moved into this level of benefit on the MADRS at 24 months—a significant increase vis-à-vis the 12-month assessment (Table 3).

Across the sample, clinical outcomes generally improved during the second year of stimulation. VNS is known to have a slow onset of clinical improvement in both epilepsy and MDD,^17,19,58,59^ often taking 6 to 18 months to manifest peak improvement. In the RECOVER trial, VNS electrical parameters were fixed after 2 months titration. It is possible that more accelerated responses could be observed if more flexible dosing was allowed in the first year. In this study, clinical outcomes at 24 months were generally superior to those at 12 months. Up to 60% of participants (median of ∼38% across measures) with NB at 12 months achieved MB at 24 months (Table 6). This finding demonstrated that a significant subset of participants with markedly TRD required more than a year to achieve meaningful benefit with VNS therapy. Extent of treatment resistance may not only predict the likelihood of acute benefit and the durability of obtained benefit, but also the intensity or duration of treatment needed to achieve benefit. For instance, the number of ECT sessions needed to achieve response has shown a sequential increase over time as the intervention has been used with more resistant patients, and longer courses of ECT (i.e., >12 sessions) are now commonly required.^60^ Similarly, there are considerable individual differences in the time to benefit with VNS, and some patients may require at least 2 years of VNS to determine their responsivity.

The findings regarding durability of benefit across multiple domains (depressive symptoms, daily function, and QoL) support the validity of the conclusions, which are consistent with previous studies of VNS in TRD.^17,18^ RECOVER is among the first of the long-term MDD treatment trials to examine durability, not only for reductions in symptom measures, but also for improvements in daily function and QoL. In general, improvements in function and QoL were equally or *slightly more durable* than improvements in symptom measures. The tripartite metric that evaluated outcomes across the 3 domains identified 80% of the sample as having meaningful benefit or better (≥MB) at 12 months, and approximately 80% of those participants maintained this level of benefit at 24 months. Thus, by this measure, approximately two-thirds of the sample benefited by 1 year and had sustained benefit during the second year of stimulation. In contrast, after inadequate response to 2 medication in the STAR*D trial, with subsequent pharmacological interventions less than 5% of patients achieved sustained remission at 1 year. The RECOVER VNS sample was likely the most resistant ever studied in a randomized controlled trial, having not benefited sufficiently from more than 13 lifetime antidepressant treatment trials, as well as interventional treatments with established efficacy in TRD (ECT, TMS, esketamine). In this context, the durability of benefit was exceptional.

Importantly, this study documented strong durability of benefit in the group of patients treated with active VNS and TAU for 2 years. It did not demonstrate that VNS was necessary in achieving this sustained benefit. Such a demonstration would require a randomized, masked, discontinuation design where patients who benefited from VNS are assigned to maintenance treatment with active or sham VNS. Given the nature of the VNS population, discontinuation studies of this type are unlikely on ethical grounds. There is documentation of a lead break followed by relapse in a patient with markedly TRD responding well to VNS, which was reversed upon lead repair.^61^

One major alternative explanation for observed durability findings could potentially be changes in concomitant treatment during the second year, during which there were no constraints on concomitant treatment. However, examination of the numbers and classes of medications added and subtracted during the second year, and specifically the receipt of ECT, TMS, and esketamine, did not yield a pattern suggesting impact on durability of benefit. These findings are consistent with the view that active VNS was an essential component to the durability of benefit.

It is not known why VNS results in strong durability in patients with markedly TRD. It has been suggested that antidepressant interventions with slower onset to clinical benefit display greater durability than faster-acting interventions, perhaps reflecting differences in neuroplastic processes.^11^ In addition, VNS, like deep brain stimulation (DBS), involves continuous neurostimulation with compliance ensured. The beneficial effects of DBS on motor symptoms of Parkinson’s disease often persist for 10 years or more, despite continued degeneration of dopamine-producing cells in the substantia nigra, and DBS in markedly TRD appears to have substantial durability.^62,63^ The strong durability of VNS in individuals with markedly TRD could be related to its slow onset of peak benefit and/or the use of continuous neurostimulation.

In summary, this study examined durability of benefit in a sample of patients with markedly TRD who were profoundly disabled, chronically ill, and had histories of inadequate antidepressant benefit from numerous trials of medication, psychotherapy, and interventional treatments. A substantial subset of participants treated with VNS and TAU achieved benefit at 12 months across measures of symptoms, function, and QoL. About 80% of those who benefited after 12 months of VNS maintained or improved upon these benefits over the ensuing year. Additionally, another subset (∼38%) of participants who did not achieve meaningful benefit at 12 months did so by the end of the second year.

## Supporting information

List of Institutional Review Boards

## Supplementary Material

Supplementary Material is available on medRxiv.

## Acknowledgments

The authors are deeply grateful to the patients and their families for participating in the RECOVER study. We greatly appreciate the Centers for Medicare & Medicaid Services for providing financial support for the Vagus Nerve Stimulation therapy system devices and implantation surgeries. The authors would also like to thank Zeeba Kabir, PhD, and Paul Cao, PhD, of Simpson Healthcare, for their editorial assistance in accordance with the Good Publication Practice 2022 guidelines. This support was funded by LivaNova.

We would also like to thank **the members of the RECOVER study group:** Advanced Mental Health Care, Palm Beach, FL (Aron Tendler); Alivation Research, Lincoln, NE (Walter Duffy); AMR Baber Research, Naperville, IL (Riaz Baber); APG Research, Orlando, FL (Morteza Nadjafi); ATP Clinical Research, Costa Mesa, CA (Gustavo Alva); Barnes-Jewish Hospital, St Louis, MO (Donald Bohnenkamp); Beacon Medical Group Behavioral Health South Bend, South Bend, IN (Suhayl Nasr); Carilion Clinic, Roanoke, VA (Anita Kablinger); Center for Anxiety and Depression, Mercer Island, WA (David Dunner); Center for Neuropsychiatry and Brain Stimulation, Durham, NC (Sandeep Vaishnavi); Charak Center for Health and Wellness, Garfield Heights, OH (Rakesh Ranjan); DENT Neurologic Institute, Amherst, NY (Horacio Capote); Emory University, Atlanta, GA (Patricio Riva Posse IV); Florida Behavioral Medicine, Largo, FL (Ashok Patel); Florida Center for TMS, Orlando, FL (Todd Broder); Florida Center for TMS, St Augustine, FL (Heather Luing); Galiz Research, Hialeah, FL (Jose Gamez); Hapworth Research, New York, NY (William Hapworth); Healthy Perspectives, Nashua, NH (Hisham Hafez); Icahn School of Medicine at Mount Sinai, New York, NY (James Murrough); Kaizen Brain Center, La Jolla, CA (Mohammed Ahmed); Marshall Psychiatry, Huntington, WV (Suzanne Holroyd); Massachusetts General Hospital, Boston, MA (Cristina Cusin); Medical College of Georgia at Augusta University, Augusta, GA (Peter Rosenquist); Medical University of South Carolina, Charleston, SC (Mark George); Michigan Clinical Research Institute, Ann Arbor, MI (Rajaprabhakaran Rajarethinam); Mindful Behavioral Health, Boca Raton, FL (Ivan Cichowicz); Neuropsychiatric Associates at Woodstock Research Center, Woodstock, VT (Susan Smiga); NeuroScience and TMS Treatment Center, Brentwood, TN (Jonathan Becker); Northwest Behavioral Research Center, Marietta, GA (Michael Banov); Nova Psychiatry, Orlando, FL (David Medina); Offices of Psychiatry & Counseling Services, Moosic, PA (Matthew Berger); Ohio State University, Columbus, OH (Kevin Reeves); OU Physicians, Tulsa, OK (Ondria Gleason); Precise Research Centers, Flowood, MS (Joseph Kwentus); PsychCare Consultants Research, St Louis, MO (Mohd Malik); Psychiatry Care and Research Center, O’Fallon, MO (John Canale); Rush University Medical Center, Chicago, IL (John Zajecka); Seattle Neuropsychiatric Treatment Center, Seattle, WA (Rebecca Allen); SF-Care, San Rafael, CA (Jason Bermak); Sheppard Pratt Health Systems, Baltimore, MD (Scott Aaronson); Signature Research Associates, Fairlawn, OH (Anand Chaturvedi); Southern Illinois University School of Medicine, Springfield, IL (Jeffrey I. Bennett); Stedman Clinical Trials, Tampa, FL (Mary Stedman); Stony Brook University Hospital, Stony Brook, NY (Lucian Manu); Syrentis Clinical Research, Santa Ana, CA (John Duffy); Texas Tech University Health Science Center, El Paso, TX (Peter Thompson); Trinity Medical, Lewiston, NY (Alfred Belen III); UC San Diego, San Diego, CA (Mounir Soliman); University of Alabama Heersink School of Medicine, Birmingham, AL (Matthew Macaluso); University of Alabama Huntsville Regional Medical Center, Huntsville, AL (Richard Shelton); University of Minnesota, Minneapolis, MN (Ziad Nahas); University of Missouri, Columbia, MO (Muaid Ithman); University of Utah Neuropsychiatric Institute, Salt Lake City, UT (Brian Mickey); University of Wisconsin, Madison, WI (Steven Garlow); UPenn Perelman School of Medicine, Philadelphia, PA (Yvette Sheline); USC Keck School of Medicine, Los Angeles, CA (Ashraf Elmashat); UT Dell Medical School, Austin, TX (Julie Farrington); and UT McGovern Medical School, Houston, TX (João Quevedo).

## Author Contributions

Charles R. Conway (Conceptualization, Investigation, Methodology, Project administration, Supervision, Validation, Visualization, Writing (original draft), Writing (review & editing)), A. John Rush (Conceptualization, Investigation, Methodology, Supervision, Validation, Visualization, Writing (original draft), Writing (review & editing)), Scott T. Aaronson (Conceptualization, Investigation, Methodology, Supervision, Validation, Visualization, Writing (original draft), Writing (review & editing)), Mark T. Bunker (Conceptualization, Methodology, Supervision, Validation, Visualization, Writing (original draft), Writing (review & editing)), Charles Gordon (Conceptualization, Data curation, Formal analysis, Methodology, Software, Validation, Visualization, Writing (original draft), Writing (review & editing)), Mark S. George (Conceptualization, Investigation, Methodology, Validation, Writing (review & editing)), Patricio Riva-Posse (Investigation, Validation, Writing (review & editing)), Rebecca M. Allen (Investigation, Validation, Writing (review & editing)), Ziad Nahas (Conceptualization, Investigation, Methodology, Validation, Writing (review & editing)), Christopher L. Kriedt (Investigation, Validation, Writing (review & editing)), John Zajecka (Investigation, Validation, Writing (review & editing)), David L. Dunner (Investigation, Validation, Writing (review & editing)), João Quevedo (Investigation, Validation, Writing (review & editing)), Yvette Sheline (Investigation, Validation, Writing (review & editing)), Walter Duffy (Investigation, Validation, Writing (review & editing)), Brian J. Mickey (Investigation, Validation, Writing (review & editing)), Mary Stedman (Investigation, Validation, Writing (review & editing)), Gustavo Alva (Investigation, Validation, Writing (review & editing)), Lucian Manu (Investigation, Validation, Writing (review & editing)), Quyen Tran (Data curation, Investigation, Project administration, Validation, Writing (review & editing)), Charles F. Zorumski (Investigation, Validation, Writing (review & editing)), Matthew Macaluso (Investigation, Validation, Writing (review & editing)), Michael Banov (Investigation, Validation, Writing (review & editing)), Cristina Cusin (Investigation, Validation, Writing (review & editing)), Jeffrey I. Bennett (Investigation, Validation, Writing (review & editing)), Hunter Brown (Investigation, Validation, Writing (review & editing)), Jeffrey Way (Data curation, Funding acquisition, Project administration, Validation, Writing (review & editing)), Olivia Shy (Data curation, Formal analysis, Software, Validation, Visualization, Writing (review & editing)), Ying-Chieh (Lisa) Lee (Data curation, Formal analysis, Software, Validation, Visualization, Writing (review & editing)), R. Hamish McAllister-Williams (Validation, Writing (review & editing)), Roger S. McIntyre (Validation, Writing (review & editing)), Harold A. Sackeim (Conceptualization, Methodology, Supervision, Validation, Visualization, Writing (original draft), Writing (review & editing))

## Data Availability Statement

The statistical analyses that are reported in the paper will be shared upon reasonable request to the corresponding author. The raw data are not publicly available.

## References

1. Rush AJ, Trivedi MH, Wisniewski SR, et al. Acute and longer-term outcomes in depressed outpatients requiring one or several treatment steps: a STAR*D report. Am J Psychiatry. 2006;163(11):1905–1917. doi:10.1176/ajp.2006.163.11.1905

2. McIntyre RS, Alsuwaidan M, Baune BT, et al. Treatment-resistant depression: definition, prevalence, detection, management, and investigational interventions. World Psychiatry. 2023;22(3):394–412. doi:10.1002/wps.21120

3. McAllister-Williams RH, Arango C, Blier P, et al. The identification, assessment and management of difficult-to-treat depression: an international consensus statement. J Affect Disord. 2020;267:264–282. doi:10.1016/j.jad.2020.02.023

4. Crown WH, Finkelstein S, Berndt ER, et al. The impact of treatment-resistant depression on health care utilization and costs. J Clin Psychiatry. 2002;63(11):963–971. doi:10.4088/jcp.v63n1102

5. Bergfeld IO, Mantione M, Figee M, Schuurman PR, Lok A, Denys D. Treatment-resistant depression and suicidality. J Affect Disord. 2018;235:362–367. doi:10.1016/j.jad.2018.04.016

6. Lundberg J, Cars T, Lööv S-Å, et al. Association of treatment-resistant depression with patient outcomes and health care resource utilization in a population-wide study. JAMA Psychiatry. 2023;80(2):167–175. doi:10.1001/jamapsychiatry.2022.3860

7. Amos TB, Tandon N, Lefebvre P, et al. Direct and indirect cost burden and change of employment status in treatment-resistant depression: a matched-cohort study using a US commercial claims database. J Clin Psychiatry. 2018;79(2):17m11725. doi:10.4088/JCP.17m11725

8. Sackeim HA, Aaronson ST, Bunker MT, et al. Update on the assessment of resistance to antidepressant treatment: Rationale for the Antidepressant Treatment History Form: Short Form-2 (ATHF-SF2). J Psychiatr Res. 2024;176:325–337. doi:10.1016/j.jpsychires.2024.05.046

9. Elmaadawi A, Naha I, Prabhudesai S, Eltohami M. Personalizing ketamine therapy: real-world predictors of response to IV ketamine and intranasal esketamine in treatment-resistant depression. Psychiatry Res. 2025;354:116821. doi:10.1016/j.psychres.2025.116821

10. Sackeim HA, Prudic J, Devanand DP, Decina P, Kerr B, Malitz S. The impact of medication resistance and continuation pharmacotherapy on relapse following response to electroconvulsive therapy in major depression. J Clin Psychopharmacol. 1990;10(2):96–104. doi:10.1097/00004714-199004000-00004

11. Sackeim HA. Acute continuation and maintenance treatment of major depressive episodes with transcranial magnetic stimulation. Brain Stimul. 2016;9(3):313–319. doi:10.1016/j.brs.2016.03.006

12. American Psychiatric Association. The Practice of ECT: Recommendations for Treatment, Training and Privileging. Second Edition. American Psychiatric Press; 2024.

13. Jelovac A, Kolshus E, McLoughlin DM. Relapse following successful electroconvulsive therapy for major depression: a meta-analysis. Neuropsychopharmacology. 2013;38(12):2467–2474. doi:10.1038/npp.2013.149

14. Sackeim HA, Haskett RF, Mulsant BH, et al. Continuation pharmacotherapy in the prevention of relapse following electroconvulsive therapy: a randomized controlled trial. JAMA. 2001;285(10):1299–1307. doi:10.1001/jama.285.10.1299

15. Kellner CH, Knapp RG, Petrides G, et al. Continuation electroconvulsive therapy vs pharmacotherapy for relapse prevention in major depression: a multisite study from the Consortium for Research in Electroconvulsive Therapy (CORE). Arch Gen Psychiatry. 2006;63(12):1337–1344. doi:10.1001/archpsyc.63.12.1337

16. Rush AJ, Marangell LB, Sackeim HA, et al. Vagus nerve stimulation for treatment-resistant depression: a randomized, controlled acute phase trial. Biol Psychiatry. 2005;58(5):347–354. doi:10.1016/j.biopsych.2005.05.025

17. Aaronson ST, Sears P, Ruvuna F, et al. A 5-year observational study of patients with treatment-resistant depression treated with vagus nerve stimulation or treatment as usual: comparison of response, remission, and suicidality. Am J Psychiatry. 2017;174(7):640–648. doi:10.1176/appi.ajp.2017.16010034

18. Sackeim HA, Brannan SK, Rush AJ, George MS, Marangell LB, Allen J. Durability of antidepressant response to vagus nerve stimulation (VNS). Int J Neuropsychopharmacol. 2007;10(6):817–826. doi:10.1017/S1461145706007425

19. Rush AJ, Sackeim HA, Marangell LB, et al. Effects of 12 months of vagus nerve stimulation in treatment-resistant depression: a naturalistic study. Biol Psychiatry. 2005;58(5):355–363. doi:10.1016/j.biopsych.2005.05.024

20. Machado-Vieira R, Baumann J, Wheeler-Castillo C, et al. The timing of antidepressant effects: a comparison of diverse pharmacological and somatic treatments. Pharmaceuticals (Basel). 2010;3(1):19–41. doi:10.3390/ph3010019

21. Nahas Z, Marangell LB, Husain MM, et al. Two-year outcome of vagus nerve stimulation (VNS) for treatment of major depressive episodes. J Clin Psychiatry. 2005;66(9):1097–1104. doi:10.4088/jcp.v66n0902

22. Rush AJ, Sackeim HA, Conway CR, et al. Clinical research challenges posed by difficult-to-treat depression. Psychol Med. 2022;52(3):419–432. doi:10.1017/S0033291721004943

23. Veal C, Tomlinson A, Cipriani A, et al. Heterogeneity of outcome measures in depression trials and the relevance of the content of outcome measures to patients: a systematic review. The Lancet Psychiatry. 2024;11(4):285–294. doi:10.1016/S2215-0366(23)00438-8

24. McCall WV, Prudic J, Olfson M, Sackeim H. Health-related quality of life following ECT in a large community sample. J Affect Disord. 2006;90(2-3):269–274. doi:10.1016/j.jad.2005.12.002

25. McCall WV, Reboussin D, Prudic J, et al. Poor health-related quality of life prior to ECT in depressed patients normalizes with sustained remission after ECT. J Affect Disord. 2013;147(1-3):107–111. doi:10.1016/j.jad.2012.10.018

26. Conway CR, Aaronson ST, Sackeim HA, et al. Clinical characteristics and treatment exposure of patients with marked treatment-resistant unipolar major depressive disorder: a RECOVER trial report. Brain Stimul. 2024;17(2):448–459. doi:10.1016/j.brs.2024.03.016

27. Zaki N, Chen LN, Lane R, et al. Safety and efficacy with esketamine in treatment-resistant depression: long-term extension study. Int J Neuropsychopharmacol. 2025;28(6): pyaf027. doi:10.1093/ijnp/pyaf027

28. Rush AJ, Kraemer HC, Sackeim HA, et al. Report by the ACNP Task Force on response and remission in major depressive disorder. Neuropsychopharmacology. 2006;31(9):1841–1853. doi:10.1038/sj.npp.1301131

29. Prien RF, Carpenter LL, Kupfer DJ. The definition and operational criteria for treatment outcome of major depressive disorder. A review of the current research literature. Arch Gen Psychiatry. 1991;48(9):796–800. doi:10.1001/archpsyc.1991.01810330020003

30. Conway CR, Kumar A, Xiong W, Bunker M, Aaronson ST, Rush AJ. Chronic vagus nerve stimulation significantly improves quality of life in treatment-resistant major depression. J Clin Psychiatry. 2018;79(5):18m12178. doi:10.4088/JCP.18m12178

31. Sackeim HA, Rush AJ, Greco T, et al. Alternative metrics for characterizing longer-term clinical outcomes in difficult-to-treat depression: I. Association with change in quality of life. Psychol Med. 2023;53(14):6511–6523. doi:10.1017/S0033291722003798

32. Aaronson ST, Sackeim HA, Jiang M, et al. Alternative metrics for characterizing longer-term clinical outcomes in difficult-to-treat depression: II. Sensitivity to treatment effects. Aust N Z J Psychiatry. 2024;58(3):250–259. doi:10.1177/00048674231209837

33. Sackeim HA, Conway CR, Aaronson ST, et al. Characterizing the effects of vagus nerve stimulation on symptom improvement in markedly treatment-resistant major depressive disorder: a RECOVER trial report. J Affect Disord. 2025;380:135–145. doi:10.1016/j.jad.2025.03.124

34. Conway CR, Olin B, Aaronson ST, et al. A prospective, multi-center randomized, controlled, blinded trial of vagus nerve stimulation for difficult to treat depression: a novel design for a novel treatment. Contemp Clin Trials. 2020;95:106066. doi:10.1016/j.cct.2020.106066

35. Conway CR, Rush AJ, Gordon C, et al. An examination of symptoms, function and quality of life as conjoint clinical outcome domains for treatment-resistant depression. J Mood Anxiety Disord. 2025;10:100121. doi:10.1016/j.xjmad.2025.100121

36. Aaronson ST, Conway CR, Gordon C, et al. Prognostic and prescriptive predictors of treatment response to adjunctive VNS therapy in major depressive disorder: a RECOVER trial report. J Clin Psychiatry. 2025;86(3):25m15850. doi:10.4088/JCP.25m15850

37. American Psychiatric Association. Diagnostic and Statistical Manual of Mental Disorders, Fifth Edition. American Psychiatric Press; 2013.

38. Montgomery SA, Asberg M. A new depression scale designed to be sensitive to change. Br J Psychiatry. 1979;134:382–389. doi:10.1192/bjp.134.4.382

39. Rush AJ, Trivedi MH, Ibrahim HM, et al. The 16-Item Quick Inventory of Depressive Symptomatology (QIDS), clinician rating (QIDS-C), and self-report (QIDS-SR): a psychometric evaluation in patients with chronic major depression. Biol Psychiatry. 2003;54(5):573–583. doi:10.1016/s0006-3223(02)01866-8

40. Guy W. Universal scales: Clinical Global Impressions. ECDEU Assessment Manual for Psychopharmacology, revised. US Department of Health, Education and Welfare publication number ADM 76-338;1976:217–222.

41. Berk M, Ng F, Dodd S, et al. The validity of the CGI severity and improvement scales as measures of clinical effectiveness suitable for routine clinical use. J Eval Clin Pract. 2008;14(6):979–983. doi:10.1111/j.1365-2753.2007.00921.x

42. Reilly Associates Health Outcomes Research. Work Productivity and Activity Impairment Questionnaire (WPAI). Accessed April 12, 2024, http://www.reillyassociates.net

43. Jha MK, Minhajuddin A, Greer TL, Carmody T, Rush AJ, Trivedi MH. Early improvement in work productivity predicts future clinical course in depressed outpatients: findings from the CO-MED trial. Am J Psychiatry. 2016;173(12):1196–1204. doi:10.1176/appi.ajp.2016.16020176

44. Reilly MC, Zbrozek AS, Dukes EM. The validity and reproducibility of a work productivity and activity impairment instrument. Pharmacoeconomics. 1993;4(5):353–365. doi:10.2165/00019053-199304050-00006

45. Rush AJ, Conway CR, Aaronson ST, et al. Effects of vagus nerve stimulation on daily function and quality of life in markedly treatment-resistant major depression: findings from a one-year, randomized, sham-controlled trial. Brain Stimul. 2025;18(3):690–700. doi:10.1016/j.brs.2024.12.1187

46. Conway CR, Aaronson ST, Sackeim HA, et al. Vagus nerve stimulation in treatment-resistant depression: a one-year, randomized, sham-controlled trial. Brain Stimul. 2025;18(3):676–689. doi:10.1016/j.brs.2024.12.1191

47. Rush AJ, South CC, Jha MK, Grannemann BD, Trivedi MH. Toward a very brief quality of life enjoyment and satisfaction questionnaire. J Affect Disord. 2019;242:87–95. doi:10.1016/j.jad.2018.08.052

48. Endicott J, Rajagopalan K, Minkwitz M, Macfadden W, Group BS. A randomized, double-blind, placebo-controlled study of quetiapine in the treatment of bipolar I and II depression: improvements in quality of life. Int Clin Psychopharmacol. 2007;22(1):29–37. doi:10.1097/01.yic.0000224797.74771.be

49. Rush AJ, Kraemer HC, Sackeim HA, et al. Report by the ACNP Task Force on response and remission in major depressive disorder. Neuropsychopharmacology. 2006;31(9):1841–1853. doi:10.1038/sj.npp.1301131

50. Norwegian Institute of Public Health. Purpose of the ATC/DDD system. WHO Collaborating Centre for Drug Statistics Methodology. Accessed August 26, 2025, https://atcddd.fhi.no/atc_ddd_methodology/purpose_of_the_atc_ddd_system/

51. Rapaport MH, Clary C, Fayyad R, Endicott J. Quality-of-life impairment in depressive and anxiety disorders. Am J Psychiatry. 2005;162(6):1171–1178. doi:10.1176/appi.ajp.162.6.1171

52. Schechter D, Endicott J, Nee J. Quality of life of ‘normal’ controls: association with lifetime history of mental illness. Psychiatry Res. 2007;152(1):45–54. doi:10.1016/j.psychres.2006.09.008

53. Forman-Hoffman VL, Flom M, Montgomery R, Robinson A. Improvements in work productivity and activity impairment among adults with anxiety or depressive symptoms participating in a relational agent-delivered digital mental health intervention. J Occup Environ Med. 2024;66(3):e99–e105. doi:10.1097/JOM.0000000000003038

54. Belter L, Cruz R, Jarecki J. Work productivity activity impairment results from the Cure SMA 2018 community update survey (P1.9-070). Neurology. 2019;92(15_supplement):P1.9-070. doi:10.1212/WNL.92.15_supplement.P1.9-070

55. Zhang W, Bansback N, Boonen A, Young A, Singh A, Anis AH. Validity of the work productivity and activity impairment questionnaire--general health version in patients with rheumatoid arthritis. Arthritis Res Ther. 2010;12(5):R177. doi:10.1186/ar3141

56. Ishak WW, Greenberg JM, Cohen RM. Predicting relapse in major depressive disorder using patient-reported outcomes of depressive symptom severity, functioning, and quality of life in the Individual Burden of Illness Index for Depression (IBI-D). J Affect Disord. 2013;151(1):59–65. doi:10.1016/j.jad.2013.05.048

57. Kraus C, Kadriu B, Lanzenberger R, Zarate Jr CA, Kasper S. Prognosis and improved outcomes in major depression: a review. Translational Psychiatry. 2019;9(1):127. doi:10.1038/s41398-019-0460-3

58. Toffa DH, Touma L, El Meskine T, Bouthillier A, Nguyen DK. Learnings from 30 years of reported efficacy and safety of vagus nerve stimulation (VNS) for epilepsy treatment: a critical review. Seizure. 2020;83:104–123. doi:10.1016/j.seizure.2020.09.027

59. Suller Marti A, Verner R, Keezer M, et al. Reduction of generalized tonic-clonic seizures following vagus nerve stimulation therapy: CORE-VNS Study 24-month follow-up. Epilepsia. 2025;66(7):2307–2314. doi:10.1111/epi.18371

60. Sackeim HA, Prudic J, Devanand DP, et al. The benefits and costs of changing treatment technique in electroconvulsive therapy due to insufficient improvement of a major depressive episode. Brain Stimul. 2020;13(5):1284–1295. doi:10.1016/j.brs.2020.06.016

61. Conway CR, Chibnall JT, Tait RC. Vagus nerve stimulation for depression: a case of a broken lead, depression relapse, revision surgery, and restoration of patient response. Brain Stimul. 2008;1(3):227–228. doi:10.1016/j.brs.2008.05.003

62. Hitti FL, Ramayya AG, McShane BJ, Yang AI, Vaughan KA, Baltuch GH. Long-term outcomes following deep brain stimulation for Parkinson’s disease. J Neurosurg. 2020;132(1):205–210. doi:10.3171/2018.8.JNS182081

63. Himes LM, Mayberg HS, Husain MM, et al. Revisiting subcallosal cingulate deep brain stimulation for depression: long-term safety and effectiveness outcomes from a pooled analysis of 172 implanted patients. Brain Stimul. 2025;18(5):1632–1640. doi:10.1016/j.brs.2025.08.017

