## Supplemental Materials for "Durability of the Benefit of Vagus Nerve Stimulation in Markedly Treatment-Resistant Major Depression: A RECOVER Trial Report"

**Table S1.** Rates of each clinical outcome at 12, 18, and 24 months across the sample

| Outcome Classification | 12 Months | 18 Months | 24 Months |
|---|---|---|---|
| | | **MADRS** | |
| N | 213 | 208 | 186 |
| ≥MB | 45.5% (97) | 50.5% (105) | 53.2% (99) |
| SB | 29.1% (62) | 32.2% (67) | 33.3% (62) |
| MB | 16.4% (35) | 18.3% (38) | 19.9% (37) |
| Remission | 16.4% (35) | 17.3% (36) | 21.0% (39) |
| NB | 54.5% (116) | 49.5% (103) | 46.8% (87) |
| | | **QIDS-C** | |
| N | 213 | 208 | 186 |
| ≥MB | 57.3% (122) | 58.2% (121) | 61.8% (115) |
| SB | 35.2% (75) | 36.5% (76) | 40.9% (76) |
| MB | 22.1% (47) | 21.6% (45) | 21.0% (39) |
| Remission | 19.7% (42) | 22.1% (46) | 24.2% (45) |
| NB | 42.7% (91) | 41.8% (87) | 38.2% (71) |
| | | **QIDS-SR** | |
| N | 213 | 208 | 186 |
| ≥MB | 60.6% (129) | 57.2% (119) | 58.6% (109) |
| SB | 38.5% (82) | 38.9% (81) | 40.9% (76) |
| MB | 22.1% (47) | 18.3% (38) | 17.7% (33) |
| Remission | 24.9% (53) | 20.7% (43) | 23.1% (43) |
| NB | 39.4% (84) | 42.8% (89) | 41.4% (77) |
| | | **CGI-I** | |
| N | 214 | 209 | 190 |
| ≥MB | 68.7% (147) | 78.5% (164) | 77.4% (147) |
| SB | 40.2% (86) | 47.4% (99) | 51.6% (98) |
| MB | 28.5% (61) | 31.1% (65) | 25.8% (49) |
| Remission | 16.8% (36) | 15.8% (33) | 22.1% (42) |
| NB | 31.3% (67) | 21.5% (45) | 22.6% (43) |
| | | **Function (WPAI Item 6)** | |
| N | 208 | 203 | 180 |
| MB | 57.2% (119) | 65.0% (132) | 60.0% (108) |
| NB | 42.8% (89) | 35.0% (71) | 40.0% (72) |
| | | **QoL (Mini-Q-LES-Q)** | |
| N | 206 | 200 | 178 |
| MB | 63.1% (130) | 67.0% (134) | 66.3% (118) |
| NB | 36.9% (76) | 33.0% (66) | 33.7% (60) |
| | | **Tripartite Metric** | |
| N | 200 | 194 | 171 |
| ≥MB | 80.0% (160) | 82.0% (159) | 81.9% (140) |

| | | | |
|---|---|---|---|
| SB | 61.0% (122) | 64.9% (126) | 65.5% (112) |
| MB | 19.0% (38) | 17.6% (33) | 19.7% (28) |
| NB | 20.0% (40) | 18.0% (35) | 18.1% (31) |

Data presented as % (n).

Abbreviations: CGI-I, Clinical Global Impression–Improvement; MADRS, Montgomery-Åsberg Depression Rating Scale; MB, meaningful benefit; Mini-Q-LES-Q, 7-item subset of the Quality of Life Enjoyment and Satisfaction Questionnaire; NB, no meaningful benefit; QIDS-C, Quick Inventory of Depressive Symptomatology–Clinician; QIDS-SR, Quick Inventory of Depressive Symptomatology–Self-Report; SB, substantial benefit; WPAI, Work Productivity and Activity Impairment Questionnaire.

**Table S2.** Proportion of participants with missing data at month 24 based on month 12 outcome

| | Participants at 12 Months | | | Missing Data at 24 Months | |
|---|---|---|---|---|---|
| **Outcome Measure** | **N** | **≥MB** | **NB** | **≥MB at 12 Months** | **NB at 12 Months** |
| **MADRS** | 221 | 97 | 124 | 15.5% | 16.1% |
| **QIDS-C** | 221 | 124 | 97 | 16.1% | 15.5% |
| **QIDS-SR** | 220 | 130 | 90 | 13.1% | 18.9% |
| **CGI-I** | 221 | 149 | 72 | 13.4% | 15.3% |
| **Function (WPAI item 6)** | 217 | 121 | 96 | 16.5% | 17.7% |
| **QoL (Mini-Q-LES-Q)** | 212 | 131 | 81 | 14.5% | 18.5% |
| **Tripartite metric** | 209 | 164 | 45 | 17.7% | 20.0% |

Abbreviations: CGI-I, Clinical Global Impression–Improvement; MADRS, Montgomery-Åsberg Depression Rating Scale; MB, meaningful benefit; Mini-Q-LES-Q, 7-item subset of the Quality of Life Enjoyment and Satisfaction Questionnaire; NB, no meaningful benefit; QIDS-C, Quick Inventory of Depressive Symptomatology–Clinician; QIDS-SR, Quick Inventory of Depressive Symptomatology–Self-Report; QoL, quality of life; WPAI, Work Productivity and Activity Impairment Questionnaire.

**Table S3.** Percentage of participants who maintained meaningful benefit or substantial benefit at 18- and 24-month follow-up

| Maintenance of ≥MB in Participants With ≥MB at 12 Months | | | | |
|---|---|---|---|---|
| **Outcome Measure (N at Month 12)** | **≥MB at 18 Months** | | **≥MB at 24 Months** | |
| | **Observed** | **Imputed** | **Observed** | **Imputed** |
| MADRS (97) | 83.0% (78/94) | 80.4% | 82.9% (68/82) | 70.1% |
| QIDS-C (122) | 81.5% (97/119) | 79.5% | 80.8% (84/104) | 68.9% |
| QIDS-SR (129) | 78.0% (99/127) | 76.7% | 79.6% (90/113) | 69.8% |
| CGI-I (147) | 89.0% (129/145) | 87.8% | 85.3% (110/129) | 74.8% |
| QoL (Mini-Q-LES-Q; 130) | 87.5% (112/128) | 86.2% | 81.3% (91/112) | 70.0% |
| Function (WPAI item 6; 119) | 83.1% (98/118) | 82.4% | 79.2% (80/101) | 67.2% |
| Tripartite metric (160) | 89.2% (140/157) | 87.5% | 89.6% (121/135) | 75.6% |
| **Maintenance of ≥MB in Participants With SB at 12 Months** | | | | |
| **Outcome Measure (N at Month 12)** | **≥MB at 18 Months** | | **≥MB at 24 Months** | |
| | **Observed** | **Imputed** | **Observed** | **Imputed** |
| MADRS (62) | 95.0% (57/60) | 91.9% | 92.6% (50/54) | 80.6% |
| QIDS-C (75) | 87.8% (65/74) | 86.7% | 90.3% (56/62) | 74.7% |
| QIDS-SR (82) | 92.6% (74/80) | 90.2% | 87.3% (62/71) | 75.6% |
| CGI-I (86) | 95.3% (81/85) | 94.2% | 92.2% (71/77) | 82.6% |
| Tripartite metric (122) | 93.3% (111/119) | 91.0% | 94.2% (97/103) | 79.5% |
| **Maintenance of SB in Participants With SB at 12 Months** | | | | |
| **Outcome Measure (N at Month 12)** | **SB at 18 Months** | | **SB at 24 Months** | |
| | **Observed** | **Imputed** | **Observed** | **Imputed** |
| MADRS (62) | 75.0% (45/60) | 72.6% | 74.1% (40/54) | 64.5% |
| QIDS-C (75) | 74.3% (55/74) | 73.3% | 79.0% (49/62) | 65.3% |
| QIDS-SR (82) | 78.8% (63/80) | 76.8% | 74.6% (53/71) | 64.6% |
| CGI-I (86) | 80.0% (68/85) | 79.1% | 83.1% (64/77) | 74.4% |
| Tripartite metric (122) | 84.9% (101/119) | 82.8% | 80.6% (83/103) | 68.0% |

Observed: Proportions were calculated using all available data at each visit.
Imputed: Proportions were calculated using the number of participants with available data at the month 12 visit as the denominator; missing data were considered as not meeting the response outcome.

Abbreviations: CGI-I, Clinical Global Impression–Improvement; MADRS, Montgomery-Åsberg Depression Rating Scale; MB, meaningful benefit; Mini-Q-LES-Q, 7-item subset of the Quality of Life Enjoyment and Satisfaction Questionnaire; QIDS-C, Quick Inventory of Depressive Symptomatology–Clinician; QIDS-SR, Quick Inventory of Depressive Symptomatology–Self-Report; QoL, quality of life; SB, substantial benefit; WPAI, Work Productivity and Activity Impairment Questionnaire.

**Table S4.** Mean (SD) concomitant psychotropic medications at month 12 and last follow-up (month 18 or 24) for the total sample, participants with meaningful benefit on the tripartite metric (score ≥1), and participants with substantial benefit on the CGI-I (score ≤2)

| | **Total Sample (N=214)** | | |
|---|---|---|---|
| | **Month 12** | **Last Follow-Up[b]** | ***P*** |
| Total Psychotropics[a] | 4.6 (2.4) | 4.6 (2.5) | 0.892 |
| Antidepressants | 1.8 (0.9) | 1.8 (0.9) | 1.000 |
| Anxiolytics | 0.8 (0.8) | 0.8 (0.8) | 0.224 |
| Antipsychotics | 0.5 (0.6) | 0.5 (0.6) | 0.533 |
| Analgesics | 0.4 (0.8) | 0.4 (0.9) | 0.096 |
| Anticonvulsants | 0.5 (0.7) | 0.5 (0.7) | 0.819 |
| Hypnotics | 0.2 (0.5) | 0.2 (0.5) | 0.083 |
| Stimulants | 0.3 (0.6) | 0.4 (0.6) | 0.286 |
| | **MB on the Tripartite Metric (n=142)** | | |
| | **Month 12** | **Last Follow-Up[b]** | ***P*** |
| Total Psychotropics[a] | 4.6 (2.5) | 4.6 (2.6) | 0.750 |
| Antidepressants | 1.8 (0.9) | 1.8 (0.9) | 1.000 |
| Anxiolytics | 0.8 (0.8) | 0.8 (0.8) | 0.452 |
| Antipsychotics | 0.4 (0.6) | 0.4 (0.6) | 0.740 |
| Analgesics | 0.4 (0.9) | 0.5 (1.0) | 0.158 |
| Anticonvulsants | 0.5 (0.7) | 0.5 (0.7) | 1.000 |
| Hypnotics | 0.2 (0.5) | 0.2 (0.5) | 0.083 |
| Stimulants | 0.3 (0.6) | 0.4 (0.6) | 0.250 |
| | **SB on the CGI-I (n=70)** | | |
| | **Month 12** | **Last Follow-Up[b]** | ***P*** |
| Total Psychotropics[a] | 4.6 (2.8) | 4.6 (3.0) | 0.816 |
| Antidepressants | 1.9 (0.8) | 1.9 (0.8) | 0.621 |
| Anxiolytics | 0.8 (0.9) | 0.8 (0.8) | 0.798 |
| Antipsychotics | 0.4 (0.6) | 0.4 (0.6) | 0.567 |
| Analgesics | 0.4 (0.9) | 0.4 (1.0) | 0.182 |
| Anticonvulsants | 0.4 (0.8) | 0.5 (0.8) | 0.321 |
| Hypnotics | 0.2 (0.5) | 0.2 (0.5) | 0.083 |
| Stimulants | 0.2 (0.5) | 0.2 (0.5) | 0.567 |

[a]Includes all categories noted above.
[b]Last follow-up is at month 24, if available, otherwise it is at month 18.

Abbreviations: CGI-I, Clinical Global Impression–Improvement; MB, meaningful benefit; SB, substantial benefit; SD, standard deviation.

**Table S5.** Proportion of participants receiving interventional treatment at any time **during the 90 days** prior to month 12 and last follow-up (month 18 or 24) for the total sample, participants with meaningful benefit on the tripartite metric (score ≥1), and participants with substantial benefit on the CGI-I (score ≤2)

| | **Total Sample (N=214)** | |
|---|---|---|
| | **Month 12** | **Last Follow-Up[a]** |
| **ECT** | 6 (2.8%) | 6 (2.8%) [1] |
| **TMS** | 2 (0.9%) | 3 (1.4%) [1] |
| **Ketamine/esketamine** | 23 (10.7%) | 24 (11.2%) [2] |
| | **MB on the Tripartite Metric (n=142)** | |
| | **Month 12** | **Last Follow-Up[a]** |
| **ECT** | 2 (1.4%) | 1 (0.7%) [0] |
| **TMS** | 1 (0.7%) | 1 (0.7%) [0] |
| **Ketamine/esketamine** | 14 (9.9%) | 15 (10.6%) [2] |
| | **SB on the CGI-I (n=70)** | |
| | **Month 12** | **Last Follow-Up[a]** |
| **ECT** | 3 (4.3%) | 2 (2.9%) [0] |
| **TMS** | 2 (2.9%) | 2 (2.9%) [0] |
| **Ketamine/esketamine** | 6 (8.6%) | 7 (10.0%) [1] |

Counts in [ ] brackets indicate participants using interventional treatment at the visit date who were not counted at month 12.

[a]Last follow-up is at month 24, if available, otherwise it is at month 18.

Abbreviations: CGI-I, Clinical Global Impression–Improvement; ECT, electroconvulsive therapy; MB, meaningful benefit; SB, substantial benefit; TMS, transcranial magnetic stimulation.

**Figure S1.** Alluvial diagram of depressive symptom measures, including (A) MADRS, (B) QIDS-C, and (C) QIDS-SR findings over 18 and 24 months

A. MADRS

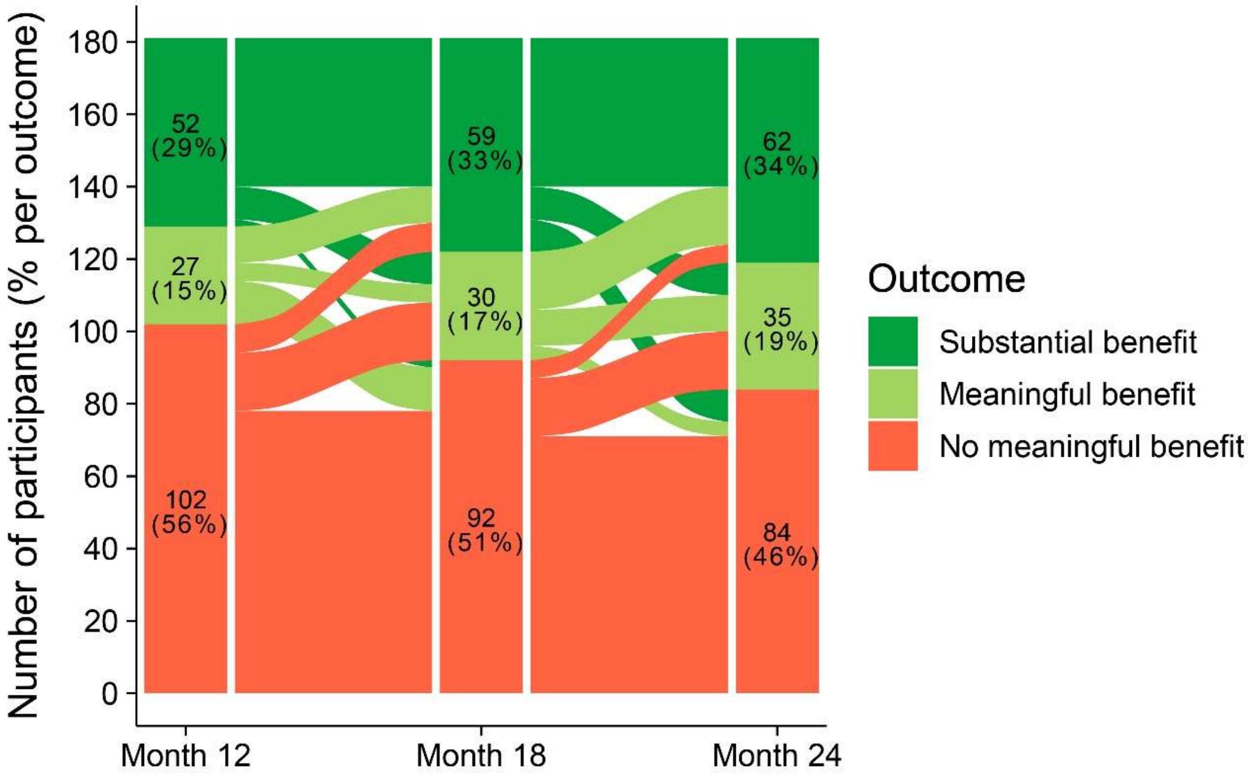

B. QIDS-C

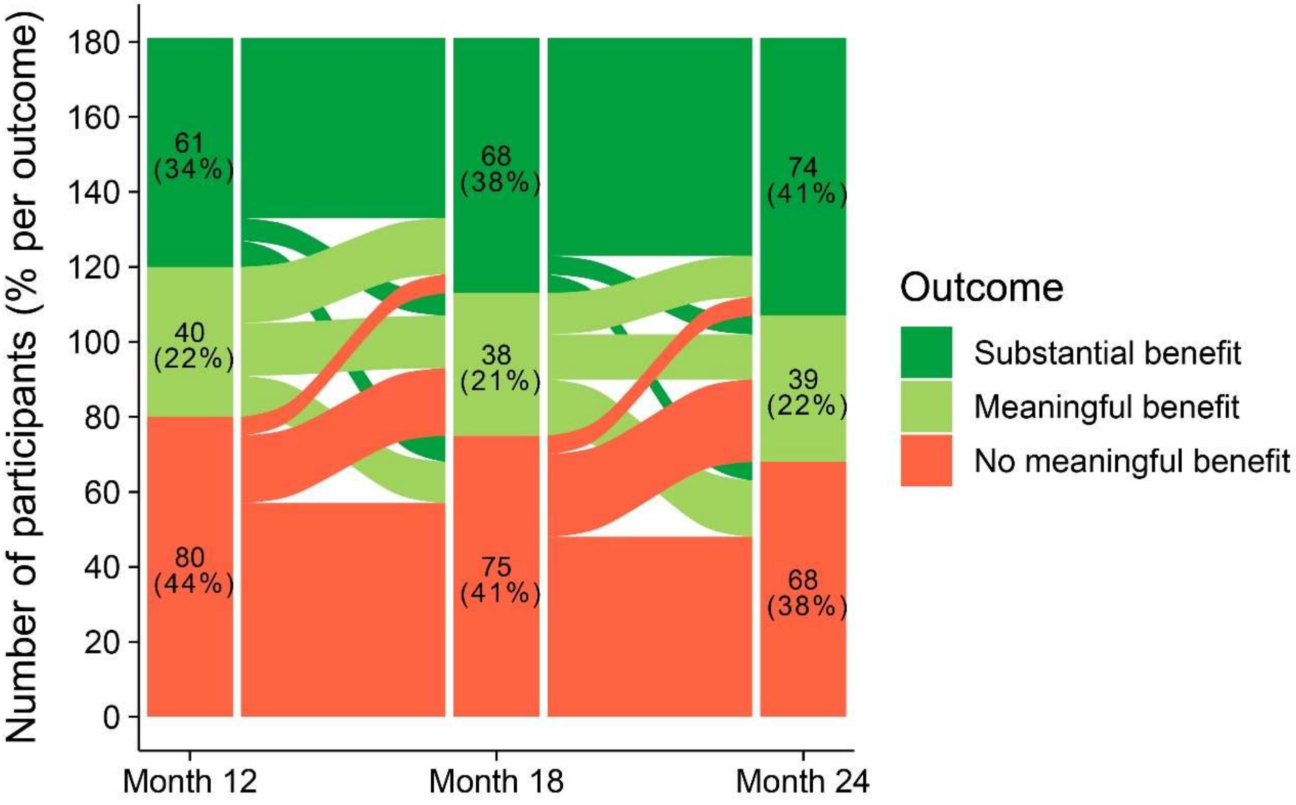
Number of participants (% per outcome)
180
160
140
120
100
80
60
40
20
0
61
(34%)
40
(22%)
80
(44%)
68
(38%)
38
(21%)
75
(41%)
74
(41%)
39
(22%)
68
(38%)
Month 12
Month 18
Month 24
Outcome
Substantial benefit
Meaningful benefit
No meaningful benefit

C. QIDS-SR

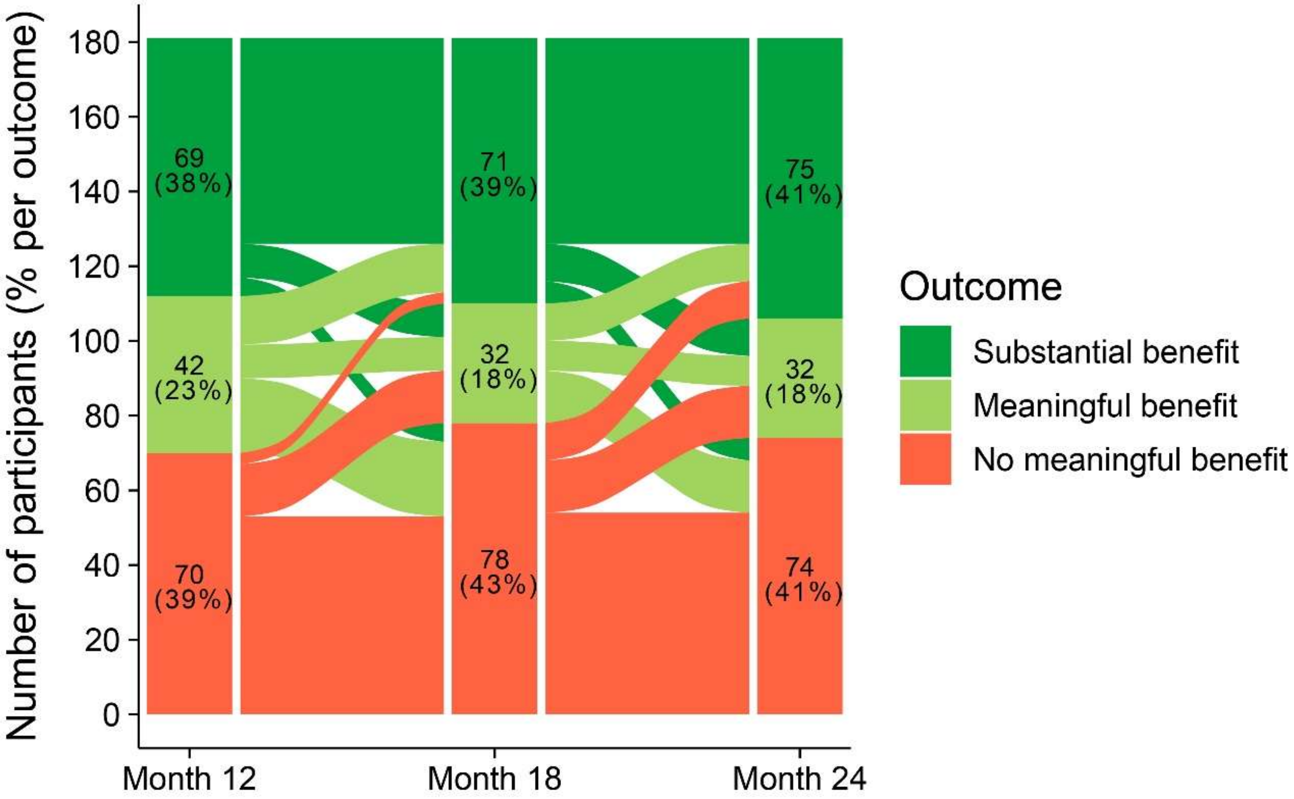

Abbreviations: MADRS, Montgomery-Åsberg Depression Rating Scale; QIDS-C, Quick Inventory of Depressive Symptomatology–Clinician; QIDS-SR, Quick Inventory of Depressive Symptomatology–Self-Report.

**Figure S2.** Alluvial diagram of (A) Mini-Q-LES-Q and (B) WPAI item 6 findings over 18 and 24 months

A. Mini-Q-LES-Q

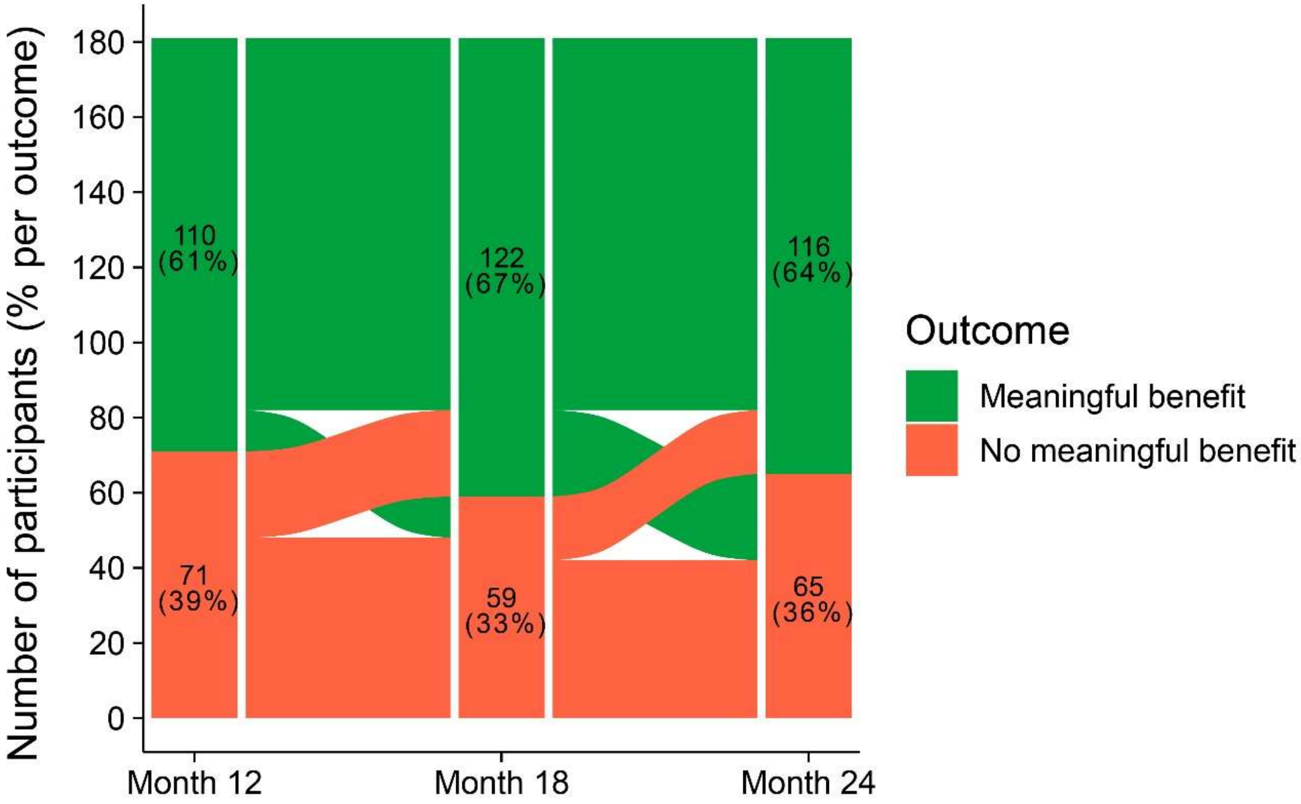

B. WPAI item 6

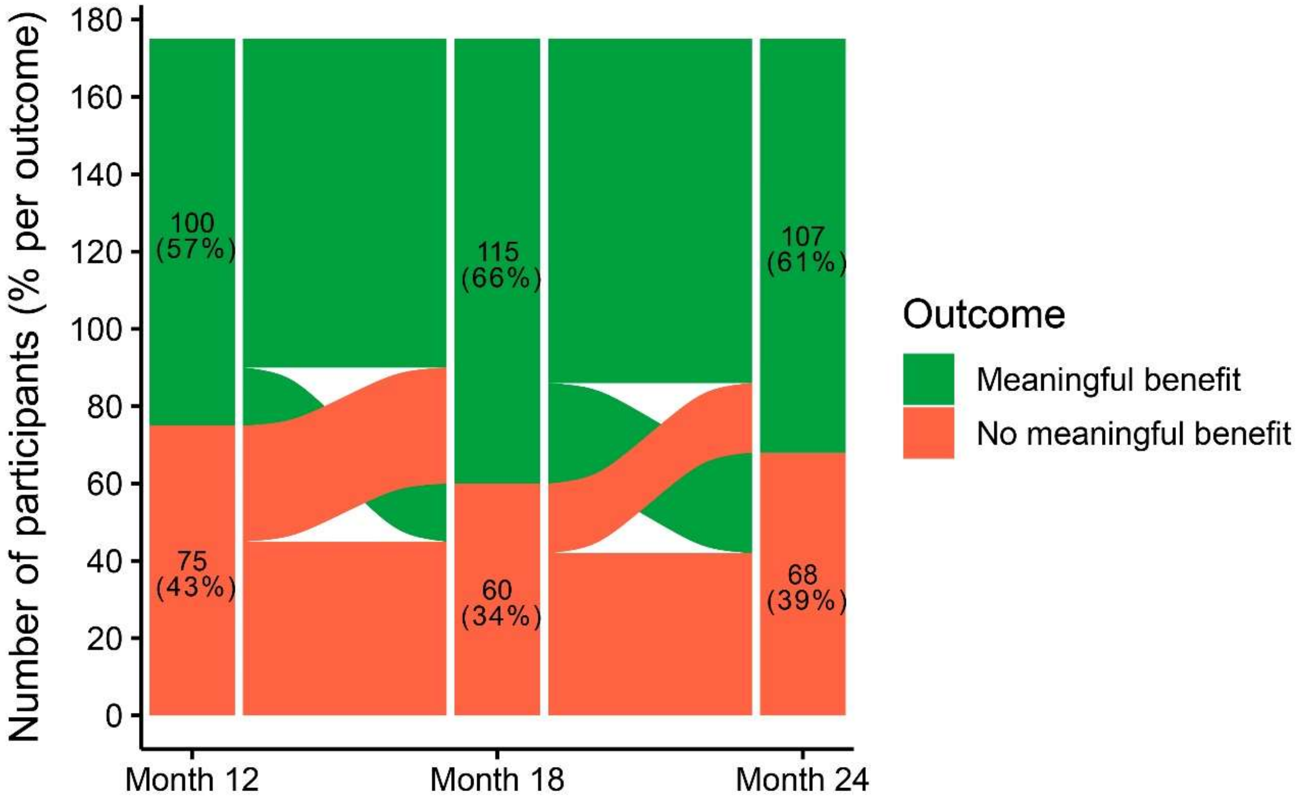

Abbreviations: Mini-Q-LES-Q, 7-item subset of the Quality of Life Enjoyment and Satisfaction Questionnaire; WPAI, Work Productivity and Activity Impairment Questionnaire.

**Supplemental Methods 1.** Identification and classification of concomitant psychotropic medications and interventional treatments in the RECOVER trial

**Concomitant Medications**. All ongoing medications were documented at each assessment occasion based on patient reporting and categorized into pharmacological classes with modification of the WHO Anatomical Therapeutic Chemical (ATC) classification system. Information on the dosage of medications was not examined in the current study. The universe of medications considered was restricted to ATC Level 1 N: Nervous system, that is, compounds with psychotropic properties. Modifications to the WHO ATC classification system reflected the consensus judgement of the 6 co-authors with expertise in clinical pharmacology, neuromodulation, and the treatment of treatment-resistant depression (TRD; SA, MB, CC, HMW, HAS, and AJR). The RECOVER database included 209 entries of medications or medication combinations categorized by the WHO ATC classification system as nervous system agents. Each entry was reviewed, and 14 that concerned agents with questionable psychotropic properties or relevance to therapeutic effects in TRD were identified. These excluded medications are listed below.

**Medications not considered psychotropics**

| **Medication** | **Indication or Use** |
|---|---|
| BETHANECHOL CHLORIDE | Postoperative and postpartum urinary retention |
| BUPIVACAINE | Anesthetic |
| CANNABIDIOL/ CANNABIS SATIVA | Treat seizures associated with Lennox-Gastaut syndrome, Dravet syndrome, or tuberous sclerosis complex |
| CEVIMELINE | Treatment for dry mouth |

| | |
|---|---|
| DIPHENHYDRAMINE/ DIPHENHYDRAMINE HYDROCHLORIDE | Antihistamine |
| EPINEPHRINE | For emergency treatment of allergic reactions, including anaphylaxis |
| GAMMA-AMINOBUTYRIC ACID; MAGNESIUM OXIDE; MATRICARIA CHAMOMILLA; MELATONIN; PASSIFLORA INCARNATA; TRYPTOPHAN, L-; VALERIANA OFFICINALIS ROOT EXTRACT; WITHANIA SOMNIFERA EXTRACT | Supplement for relaxation |
| LIDOCAINE | Local anesthetic |
| METOPROLOL | Treatment for hypertension |
| PILOCARPINE | Treatment for dry mouth |
| PYRIDOSTIGMINE | Treatment for myasthenia gravis |
| THIOCTIC ACID | Treatment for peripheral diabetic polyneuropathy |
| TIZANIDINE | Management of spasticity |
| WITHANIA SOMNIFERA | Supplement for stress and anxiety, sleep, male infertility, and athletic performance |

The WHO ATC classification system further categorizes medications by chemical, pharmacological, or therapeutic subgroup (eg, analgesics, antidepressants). However, the same medication may receive multiple classifications based on differing intended uses. For example, in the RECOVER database, there were 3 entries for amitriptyline, which was classified as an analgesic, antidepressant, and anxiolytic. The statistical analyses required that each medication be assigned to 1 class so that additions and subtractions can be counted only once per medication. Therefore, consensus was also achieved on the class assigned to each medication. These classifications reflected widely accepted pharmacological distinctions. For example, amitriptyline was coded as an antidepressant, and valproic acid as an anticonvulsant (antiepileptic). Since the WHO ACT classification system does not contain a category for mood stabilizers, lithium was placed in the antidepressant class in place of its WHO assignment as an antipsychotic. The WHO class of anesthetics was removed, as 2 of 3 entries pertained to local anesthetics and the third to ketamine, which was reclassified in the antidepressant grouping.

The medication classes derived from the WHO ACT classification system are listed below. Due to their infrequency of use and limited relevance for therapeutic effects in TRD, the classes of antidementia drugs, anti-Parkinson drugs, and other nervous system drugs (for addictive disorders or miscellaneous) were combined into a single class, termed “other.” The total number of psychotropic medications at an assessment occasion corresponded to the sum of number of medications in the classes of analgesics, anticonvulsants, antidepressants, antipsychotics, anxiolytics, hypnotics, stimulants, and other.

**WHO medication classes and corresponding RECOVER classes**

| WHO ACT Class | RECOVER Class |
| --- | --- |
| Analgesics | Analgesics |
| Antidementia drugs | Other |
| Anti-Parkinson drugs | Other |
| Antidepressants | Antidepressants |
| Antiepileptics | Anticonvulsants |
| Antipsychotics | Antipsychotics |
| Anxiolytics | Anxiolytics |
| Hypnotics and sedatives | Hypnotics |
| Other nervous system drugs used in addictive disorders | Other |
| Other nervous system drugs miscellaneous | Other |
| Psychostimulants, agents used for ADHD, and nootropics | Stimulants |

The complete list of psychotropic medication entries in RECOVER and their classifications are presented in Supplemental Methods 2.

**Concomitant Interventional Psychiatry Treatments**

At each assessment occasion, ongoing treatment with esketamine or ketamine, electroconvulsive therapy (ECT), or transcranial magnetic stimulation (TMS) was also documented. There is substantial evidence regarding the efficacy of these interventions, specifically in treatment of TRD. Previous research demonstrated in the RECOVER sample that a history of treatment with ECT or TMS was prognostic of outcomes after the first year. Thus, determination of whether the use of interventional psychiatry treatments changed over the second year of RECOVER is consequential in interpreting the durability findings.

After completion of the first year's blinded, randomized phase, there was no restriction on concomitant treatments. These interventional treatments could be administered as rescue interventions, continuation/maintenance treatment, or acute courses. This study identified the number of participants receiving each type of intervention (ketamine, ECT, and/or TMS) at the month-12 and last follow-up time points, and the number of patients who added or subtracted these interventions at end point relative to month 12.

**Supplemental Methods 2. The complete list of psychotropic medication entries in RECOVER and their classifications**

| Standardized Medication Name | WHO Drug ATC Level 1 | WHO Drug ATC Level 2 | WHO Drug ATC Level 3 | **Classification in RECOVER** |
|---|---|---|---|---|
| ACETAZOLAMIDE | NERVOUS SYSTEM | ANTIEPILEPTICS | ANTIEPILEPTICS | ANTIEPILEPTICS |
| ACETYLSALICYLIC ACID | NERVOUS SYSTEM | ANALGESICS | OTHER ANALGESICS AND ANTIPYRETICS | ANALGESICS |
| ACETYLSALICYLIC ACID;CAFFEINE;PARACETAMOL | NERVOUS SYSTEM | ANALGESICS | OTHER ANALGESICS AND ANTIPYRETICS | ANALGESICS |
| ACETYLSALICYLIC ACID;CHLORPHENAMINE;PHENYLEPHRINE | NERVOUS SYSTEM | ANALGESICS | OTHER ANALGESICS AND ANTIPYRETICS | ANALGESICS |
| AGOMELATINE | NERVOUS SYSTEM | PSYCHOANALEPTICS | ANTIDEPRESSANTS | ANTIDEPRESSANTS |
| ALPRAZOLAM | NERVOUS SYSTEM | PSYCHOLEPTICS | ANXIOLYTICS | ANXIOLYTICS |
| AMANTADINE | NERVOUS SYSTEM | ANTI-PARKINSON DRUGS | DOPAMINERGIC AGENTS | OTHER |
| AMITRIPTYLINE | NERVOUS SYSTEM | PSYCHOANALEPTICS | ANTIDEPRESSANTS | ANTIDEPRESSANTS |
| AMOXAPINE | NERVOUS SYSTEM | PSYCHOANALEPTICS | ANTIDEPRESSANTS | ANTIDEPRESSANTS |
| AMPHETAMINE | NERVOUS SYSTEM | PSYCHOANALEPTICS | PSYCHOSTIMULANTS, AGENTS USED FOR ADHD AND NOOTROPICS | STIMULANTS |
| AMPHETAMINE;DEXAMPHETAMINE | NERVOUS SYSTEM | PSYCHOANALEPTICS | PSYCHOSTIMULANTS, AGENTS USED FOR ADHD AND NOOTROPICS | STIMULANTS |
| ARIPIPRAZOLE | NERVOUS SYSTEM | PSYCHOLEPTICS | ANTIPSYCHOTICS | ANTIPSYCHOTICS |
| ARMODAFINIL | NERVOUS SYSTEM | PSYCHOANALEPTICS | PSYCHOSTIMULANTS, AGENTS USED FOR ADHD AND NOOTROPICS | STIMULANTS |
| ASENAPINE | NERVOUS SYSTEM | PSYCHOLEPTICS | ANTIPSYCHOTICS | ANTIPSYCHOTICS |
| ATOGEPANT | NERVOUS SYSTEM | ANALGESICS | ANTIMIGRAINE PREPARATIONS | ANALGESICS |
| ATOMOXETINE | NERVOUS SYSTEM | PSYCHOANALEPTICS | PSYCHOSTIMULANTS, AGENTS USED FOR ADHD AND NOOTROPICS | STIMULANTS |
| BACLOFEN;GABAPENTIN;LIDOCAINE | NERVOUS SYSTEM | ANALGESICS | OTHER ANALGESICS AND ANTIPYRETICS | ANALGESICS |
| BENZATROPINE | NERVOUS SYSTEM | ANTI-PARKINSON DRUGS | ANTICHOLINERGIC AGENTS | OTHER |
| BOTULINUM TOXIN TYPE A | NERVOUS SYSTEM | ANALGESICS | ANTIMIGRAINE PREPARATIONS | ANALGESICS |
| BREXPIPRAZOLE | NERVOUS SYSTEM | PSYCHOLEPTICS | ANTIPSYCHOTICS | ANTIPSYCHOTICS |
| BUPRENORPHINE | NERVOUS SYSTEM | ANALGESICS | DRUGS USED IN ADDICTIVE DISORDERS | OTHER |
| BUPRENORPHINE;NALOXONE | NERVOUS SYSTEM | OTHER NERVOUS SYSTEM DRUGS | DRUGS USED IN ADDICTIVE DISORDERS | OTHER |
| BUPROPION | NERVOUS SYSTEM | PSYCHOANALEPTICS | ANTIDEPRESSANTS | ANTIDEPRESSANTS |
| BUPROPION;DEXTROMETHORPHAN | NERVOUS SYSTEM | PSYCHOANALEPTICS | ANTIDEPRESSANTS | ANTIDEPRESSANTS |
| BUSPIRONE | NERVOUS SYSTEM | PSYCHOLEPTICS | ANXIOLYTICS | ANXIOLYTICS |
| BUTALBITAL;CAFFEINE;PARACETAMOL | NERVOUS SYSTEM | ANALGESICS | OTHER ANALGESICS AND ANTIPYRETICS | ANALGESICS |
| BUTALBITAL;PARACETAMOL | NERVOUS SYSTEM | ANALGESICS | OTHER ANALGESICS AND ANTIPYRETICS | ANALGESICS |

| | | | | |
|---|---|---|---|---|
| CAFFEINE;CODEINE;PARACETAMOL | NERVOUS SYSTEM | ANALGESICS | OPIOIDS | ANALGESICS |
| CARBAMAZEPINE | NERVOUS SYSTEM | PSYCHOLEPTICS | ANTIPSYCHOTICS | ANTIEPILEPTICS |
| CARBIDOPA MONOHYDRATE;LEVODOPA | NERVOUS SYSTEM | ANTI-PARKINSON DRUGS | DOPAMINERGIC AGENTS | OTHER |
| CARIPRAZINE | NERVOUS SYSTEM | PSYCHOLEPTICS | ANTIPSYCHOTICS | ANTIPSYCHOTICS |
| CHLORDIAZEPOXIDE | NERVOUS SYSTEM | PSYCHOLEPTICS | ANXIOLYTICS | ANXIOLYTICS |
| CHLORPROMAZINE | NERVOUS SYSTEM | PSYCHOLEPTICS | ANTIPSYCHOTICS | ANTIPSYCHOTICS |
| CITALOPRAM | NERVOUS SYSTEM | PSYCHOANALEPTICS | ANTIDEPRESSANTS | ANTIDEPRESSANTS |
| CLOMIPRAMINE | NERVOUS SYSTEM | PSYCHOANALEPTICS | ANTIDEPRESSANTS | ANTIDEPRESSANTS |
| CLONAZEPAM | NERVOUS SYSTEM | PSYCHOLEPTICS | ANXIOLYTICS | ANXIOLYTICS |
| CLONIDINE | NERVOUS SYSTEM | PSYCHOANALEPTICS | PSYCHOSTIMULANTS, AGENTS USED FOR ADHD AND NOOTROPICS | OTHER |
| CLORAZEPIC ACID | NERVOUS SYSTEM | PSYCHOLEPTICS | ANXIOLYTICS | ANXIOLYTICS |
| CLOZAPINE | NERVOUS SYSTEM | PSYCHOLEPTICS | ANTIPSYCHOTICS | ANTIPSYCHOTICS |
| CODEINE | NERVOUS SYSTEM | ANALGESICS | OPIOIDS | ANALGESICS |
| DARIDOREXANT | NERVOUS SYSTEM | PSYCHOLEPTICS | HYPNOTICS AND SEDATIVES | HYPNOTICS |
| DESIPRAMINE | NERVOUS SYSTEM | PSYCHOANALEPTICS | ANTIDEPRESSANTS | ANTIDEPRESSANTS |
| DESVENLAFAXINE | NERVOUS SYSTEM | PSYCHOANALEPTICS | ANTIDEPRESSANTS | ANTIDEPRESSANTS |
| DEUTETRABENAZINE | NERVOUS SYSTEM | OTHER NERVOUS SYSTEM DRUGS | OTHER NERVOUS SYSTEM DRUGS | OTHER |
| DEXAMFETAMINE | NERVOUS SYSTEM | PSYCHOANALEPTICS | PSYCHOSTIMULANTS, AGENTS USED FOR ADHD AND NOOTROPICS | STIMULANTS |
| DEXMETHYLPHENIDATE | NERVOUS SYSTEM | PSYCHOANALEPTICS | PSYCHOSTIMULANTS, AGENTS USED FOR ADHD AND NOOTROPICS | STIMULANTS |
| DEXTROMETHORPHAN;QUINIDINE | NERVOUS SYSTEM | OTHER NERVOUS SYSTEM DRUGS | OTHER NERVOUS SYSTEM DRUGS | OTHER |
| DEXTROMETHORPHAN/DEXTROMETHORPHAN HYDROBROMIDE;DOXYLAMINE SUCCINATE;EPHEDRINE SULFATE;ETHANOL;PARACETAMOL/DEXTROMETHORPHAN;DOXYLAMINE;EPHEDRINE;ETHANOL;PARACETAMOL/DEXTROMETHORPHAN;DOXYLAMINE;PARACETAMOL/DEXTROMETHORPHAN;GUAIFENESIN;PARACETAMOL;PHENYLEPHRINE/DEXTROMETHORPHAN;GUAIFENESIN;PARACETAMOL;PSEUDOEPHEDRINE/DEXTROMETHORPHAN;PARACETAMOL;PHENYLEPHRINE | NERVOUS SYSTEM | ANALGESICS | OTHER ANALGESICS AND ANTIPYRETICS | ANALGESICS |
| DIAZEPAM | NERVOUS SYSTEM | PSYCHOLEPTICS | ANXIOLYTICS | ANXIOLYTICS |

| | | | | |
|---|---|---|---|---|
| DICHLORALPHENAZONE;ISOMETHEPTENE;PARACETAMOL | NERVOUS SYSTEM | ANALGESICS | OTHER ANALGESICS AND ANTIPYRETICS | ANALGESICS |
| DIMENHYDRINATE | NERVOUS SYSTEM | OTHER NERVOUS SYSTEM DRUGS | ANTIVERTIGO PREPARATIONS | OTHER |
| DISULFIRAM | NERVOUS SYSTEM | OTHER NERVOUS SYSTEM DRUGS | DRUGS USED IN ADDICTIVE DISORDERS | OTHER |
| DONEPEZIL | NERVOUS SYSTEM | PSYCHOANALEPTICS | ANTI-DEMENTIA DRUGS | OTHER |
| DONEPEZIL HYDROCHLORIDE;MEMANTINE HYDROCHLORIDE | NERVOUS SYSTEM | PSYCHOANALEPTICS | ANTI-DEMENTIA DRUGS | OTHER |
| DOXAZOSIN | NERVOUS SYSTEM | PSYCHOLEPTICS | ANXIOLYTICS | ANXIOLYTICS |
| DOXEPIN | NERVOUS SYSTEM | PSYCHOLEPTICS | HYPNOTICS AND SEDATIVES | ANTIDEPRESSANTS |
| DOXYLAMINE | NERVOUS SYSTEM | PSYCHOLEPTICS | HYPNOTICS AND SEDATIVES | HYPNOTICS |
| DULOXETINE | NERVOUS SYSTEM | PSYCHOANALEPTICS | ANTIDEPRESSANTS | ANTIDEPRESSANTS |
| ELETRIPTAN | NERVOUS SYSTEM | ANALGESICS | ANTIMIGRAINE PREPARATIONS | ANALGESICS |
| EPTINEZUMAB | NERVOUS SYSTEM | ANALGESICS | ANTIMIGRAINE PREPARATIONS | ANALGESICS |
| ERENUMAB | NERVOUS SYSTEM | ANALGESICS | ANTIMIGRAINE PREPARATIONS | ANALGESICS |
| ESCITALOPRAM | NERVOUS SYSTEM | PSYCHOANALEPTICS | ANTIDEPRESSANTS | ANTIDEPRESSANTS |
| ESKETAMINE | NERVOUS SYSTEM | PSYCHOANALEPTICS | ANTIDEPRESSANTS | ANTIDEPRESSANTS |
| ESTAZOLAM | NERVOUS SYSTEM | PSYCHOLEPTICS | HYPNOTICS AND SEDATIVES | HYPNOTICS |
| ESZOPICLONE | NERVOUS SYSTEM | PSYCHOLEPTICS | HYPNOTICS AND SEDATIVES | HYPNOTICS |
| FENTANYL | NERVOUS SYSTEM | ANALGESICS | OPIOIDS | ANALGESICS |
| FLUOXETINE | NERVOUS SYSTEM | PSYCHOANALEPTICS | ANTIDEPRESSANTS | ANTIDEPRESSANTS |
| FLUPHENAZINE | NERVOUS SYSTEM | PSYCHOLEPTICS | ANTIPSYCHOTICS | ANTIPSYCHOTICS |
| FLURAZEPAM | NERVOUS SYSTEM | PSYCHOLEPTICS | HYPNOTICS AND SEDATIVES | HYPNOTICS |
| FLUVOXAMINE | NERVOUS SYSTEM | PSYCHOANALEPTICS | ANTIDEPRESSANTS | ANTIDEPRESSANTS |
| FREMANEZUMAB VFRM | NERVOUS SYSTEM | ANALGESICS | ANTIMIGRAINE PREPARATIONS | ANALGESICS |
| GABAPENTIN | NERVOUS SYSTEM | ANTIEPILEPTICS | ANTIEPILEPTICS | ANTIEPILEPTICS |
| GALCANEZUMAB | NERVOUS SYSTEM | ANALGESICS | ANTIMIGRAINE PREPARATIONS | ANALGESICS |
| GUANFACINE | NERVOUS SYSTEM | PSYCHOANALEPTICS | PSYCHOSTIMULANTS, AGENTS USED FOR ADHD AND NOOTROPICS | STIMULANTS |
| HALOPERIDOL | NERVOUS SYSTEM | PSYCHOLEPTICS | ANTIPSYCHOTICS | ANTIPSYCHOTICS |
| HYDROCODONE | NERVOUS SYSTEM | ANALGESICS | OPIOIDS | ANALGESICS |
| HYDROCODONE;PARACETAMOL | NERVOUS SYSTEM | ANALGESICS | OPIOIDS | ANALGESICS |
| HYDROMORPHONE | NERVOUS SYSTEM | ANALGESICS | OPIOIDS | ANALGESICS |
| HYDROXYZINE | NERVOUS SYSTEM | PSYCHOLEPTICS | ANXIOLYTICS | ANXIOLYTICS |
| IMIPRAMINE | NERVOUS SYSTEM | PSYCHOANALEPTICS | ANTIDEPRESSANTS | ANTIDEPRESSANTS |
| ISOCARBOXAZID | NERVOUS SYSTEM | PSYCHOANALEPTICS | ANTIDEPRESSANTS | ANTIDEPRESSANTS |

| | | | | |
|---|---|---|---|---|
| KETAMINE | NERVOUS SYSTEM | PSYCHOANALEPTICS | ANTIDEPRESSANTS | ANTIDEPRESSANTS |
| LACOSAMIDE | NERVOUS SYSTEM | ANTIEPILEPTICS | ANTIEPILEPTICS | ANTIEPILEPTICS |
| LAMOTRIGINE | NERVOUS SYSTEM | ANTIEPILEPTICS | ANTIEPILEPTICS | ANTIEPILEPTICS |
| LASMIDITAN | NERVOUS SYSTEM | ANALGESICS | ANTIMIGRAINE PREPARATIONS | ANALGESICS |
| LEMBOREXANT | NERVOUS SYSTEM | PSYCHOLEPTICS | HYPNOTICS AND SEDATIVES | HYPNOTICS |
| LEVETIRACETAM | NERVOUS SYSTEM | ANTIEPILEPTICS | ANTIEPILEPTICS | ANTIEPILEPTICS |
| LEVOMILNACIPRAN | NERVOUS SYSTEM | PSYCHOANALEPTICS | ANTIDEPRESSANTS | ANTIDEPRESSANTS |
| LISDEXAMFETAMINE | NERVOUS SYSTEM | PSYCHOANALEPTICS | PSYCHOSTIMULANTS, AGENTS USED FOR ADHD AND NOOTROPICS | STIMULANTS |
| LITHIUM | NERVOUS SYSTEM | PSYCHOLEPTICS | ANTIPSYCHOTICS | ANTIDEPRESSANTS |
| LORAZEPAM | NERVOUS SYSTEM | PSYCHOLEPTICS | ANXIOLYTICS | ANXIOLYTICS |
| LUMATEPERONE | NERVOUS SYSTEM | PSYCHOLEPTICS | ANTIPSYCHOTICS | ANTIPSYCHOTICS |
| LURASIDONE | NERVOUS SYSTEM | PSYCHOLEPTICS | ANTIPSYCHOTICS | ANTIPSYCHOTICS |
| MECLOZINE | NERVOUS SYSTEM | OTHER NERVOUS SYSTEM DRUGS | ANTIVERTIGO PREPARATIONS | OTHER |
| MELATONIN | NERVOUS SYSTEM | PSYCHOLEPTICS | HYPNOTICS AND SEDATIVES | HYPNOTICS |
| MEMANTINE | NERVOUS SYSTEM | PSYCHOANALEPTICS | ANTI-DEMENTIA DRUGS | OTHER |
| METHADONE | NERVOUS SYSTEM | ANALGESICS | OPIOIDS | OTHER |
| METHYLPHENIDATE | NERVOUS SYSTEM | PSYCHOANALEPTICS | PSYCHOSTIMULANTS, AGENTS USED FOR ADHD AND NOOTROPICS | STIMULANTS |
| MIDAZOLAM | NERVOUS SYSTEM | PSYCHOLEPTICS | HYPNOTICS AND SEDATIVES | HYPNOTICS |
| MILNACIPRAN | NERVOUS SYSTEM | PSYCHOANALEPTICS | ANTIDEPRESSANTS | ANTIDEPRESSANTS |
| MIRTAZAPINE | NERVOUS SYSTEM | PSYCHOANALEPTICS | ANTIDEPRESSANTS | ANTIDEPRESSANTS |
| MODAFINIL | NERVOUS SYSTEM | PSYCHOANALEPTICS | PSYCHOSTIMULANTS, AGENTS USED FOR ADHD AND NOOTROPICS | STIMULANTS |
| MORPHINE | NERVOUS SYSTEM | ANALGESICS | OPIOIDS | ANALGESICS |
| NALTREXONE | NERVOUS SYSTEM | OTHER NERVOUS SYSTEM DRUGS | DRUGS USED IN ADDICTIVE DISORDERS | OTHER |
| NARATRIPTAN | NERVOUS SYSTEM | ANALGESICS | ANTIMIGRAINE PREPARATIONS | ANALGESICS |
| NEFAZODONE | NERVOUS SYSTEM | PSYCHOANALEPTICS | ANTIDEPRESSANTS | ANTIDEPRESSANTS |
| NICOTINE POLACRILEX | NERVOUS SYSTEM | OTHER NERVOUS SYSTEM DRUGS | DRUGS USED IN ADDICTIVE DISORDERS | OTHER |
| NORTRIPTYLINE | NERVOUS SYSTEM | PSYCHOANALEPTICS | ANTIDEPRESSANTS | ANTIDEPRESSANTS |
| OLANZAPINE | NERVOUS SYSTEM | PSYCHOLEPTICS | ANTIPSYCHOTICS | ANTIPSYCHOTICS |
| OLANZAPINE;SAMIDORPHAN L-MALATE | NERVOUS SYSTEM | PSYCHOLEPTICS | ANTIPSYCHOTICS | ANTIPSYCHOTICS |
| OXCARBAZEPINE | NERVOUS SYSTEM | ANTIEPILEPTICS | ANTIEPILEPTICS | ANTIEPILEPTICS |
| OXITRIPTAN | NERVOUS SYSTEM | PSYCHOANALEPTICS | ANTIDEPRESSANTS | ANTIDEPRESSANTS |
| OXYCODONE | NERVOUS SYSTEM | ANALGESICS | OPIOIDS | ANALGESICS |

| | | | | |
|---|---|---|---|---|
| OXYCODONE;PARACETAMOL | NERVOUS SYSTEM | ANALGESICS | OPIOIDS | ANALGESICS |
| PALIPERIDONE | NERVOUS SYSTEM | PSYCHOLEPTICS | ANTIPSYCHOTICS | ANTIPSYCHOTICS |
| PARACETAMOL | NERVOUS SYSTEM | ANALGESICS | OTHER ANALGESICS AND ANTIPYRETICS | ANALGESICS |
| PARACETAMOL;PHENYLEPHRINE | NERVOUS SYSTEM | ANALGESICS | OTHER ANALGESICS AND ANTIPYRETICS | ANALGESICS |
| PAROXETINE | NERVOUS SYSTEM | PSYCHOANALEPTICS | ANTIDEPRESSANTS | ANTIDEPRESSANTS |
| PERPHENAZINE | NERVOUS SYSTEM | PSYCHOLEPTICS | ANTIPSYCHOTICS | ANTIPSYCHOTICS |
| PHENELZINE | NERVOUS SYSTEM | PSYCHOANALEPTICS | ANTIDEPRESSANTS | ANTIDEPRESSANTS |
| PRAMIPEXOLE | NERVOUS SYSTEM | ANTI-PARKINSON DRUGS | DOPAMINERGIC AGENTS | OTHER |
| PRAZOSIN | NERVOUS SYSTEM | PSYCHOLEPTICS | ANXIOLYTICS | ANXIOLYTICS |
| PREGABALIN | NERVOUS SYSTEM | PSYCHOLEPTICS | ANXIOLYTICS | ANTIEPILEPTICS |
| PRIMIDONE | NERVOUS SYSTEM | ANTIEPILEPTICS | ANTIEPILEPTICS | ANTIEPILEPTICS |
| PROPRANOLOL | NERVOUS SYSTEM | PSYCHOLEPTICS | ANXIOLYTICS | ANXIOLYTICS |
| PROTRIPTYLINE | NERVOUS SYSTEM | PSYCHOANALEPTICS | ANTIDEPRESSANTS | ANTIDEPRESSANTS |
| QUETIAPINE | NERVOUS SYSTEM | PSYCHOLEPTICS | ANTIPSYCHOTICS | ANTIPSYCHOTICS |
| RAMELTEON | NERVOUS SYSTEM | PSYCHOLEPTICS | HYPNOTICS AND SEDATIVES | HYPNOTICS |
| RIMEGEPANT SULFATE | NERVOUS SYSTEM | ANALGESICS | ANTIMIGRAINE PREPARATIONS | ANALGESICS |
| RISPERIDONE | NERVOUS SYSTEM | PSYCHOLEPTICS | ANTIPSYCHOTICS | ANTIPSYCHOTICS |
| RIZATRIPTAN | NERVOUS SYSTEM | ANALGESICS | ANTIMIGRAINE PREPARATIONS | ANALGESICS |
| ROPINIROLE | NERVOUS SYSTEM | ANTI-PARKINSON DRUGS | DOPAMINERGIC AGENTS | OTHER |
| SALSALATE | NERVOUS SYSTEM | ANALGESICS | OTHER ANALGESICS AND ANTIPYRETICS | ANALGESICS |
| SELEGILINE | NERVOUS SYSTEM | PSYCHOANALEPTICS | ANTIDEPRESSANTS | ANTIDEPRESSANTS |
| SERDEXMETHYLPHENIDATE CHLORIDE | NERVOUS SYSTEM | PSYCHOANALEPTICS | PSYCHOSTIMULANTS, AGENTS USED FOR ADHD AND NOOTROPICS | STIMULANTS |
| SERTRALINE/SERTRALINE HYDROCHLORIDE | NERVOUS SYSTEM | PSYCHOANALEPTICS | ANTIDEPRESSANTS | ANTIDEPRESSANTS |
| SOLRIAMFETOL | NERVOUS SYSTEM | PSYCHOANALEPTICS | PSYCHOSTIMULANTS, AGENTS USED FOR ADHD AND NOOTROPICS | STIMULANTS |
| SUMATRIPTAN | NERVOUS SYSTEM | ANALGESICS | ANTIMIGRAINE PREPARATIONS | ANALGESICS |
| SUVOREXANT | NERVOUS SYSTEM | PSYCHOLEPTICS | HYPNOTICS AND SEDATIVES | HYPNOTICS |
| TAPENTADOL | NERVOUS SYSTEM | ANALGESICS | OPIOIDS | ANALGESICS |
| TEMAZEPAM | NERVOUS SYSTEM | PSYCHOLEPTICS | HYPNOTICS AND SEDATIVES | HYPNOTICS |
| THIORIDAZINE | NERVOUS SYSTEM | PSYCHOLEPTICS | ANTIPSYCHOTICS | ANTIPSYCHOTICS |
| TOPIRAMATE | NERVOUS SYSTEM | ANTIEPILEPTICS | ANTIEPILEPTICS | ANTIEPILEPTICS |
| TRAMADOL | NERVOUS SYSTEM | ANALGESICS | OPIOIDS | ANALGESICS |
| TRANYLCYPROMINE | NERVOUS SYSTEM | PSYCHOANALEPTICS | ANTIDEPRESSANTS | ANTIDEPRESSANTS |
| TRAZODONE | NERVOUS SYSTEM | PSYCHOANALEPTICS | ANTIDEPRESSANTS | ANTIDEPRESSANTS |

| | | | | |
|---|---|---|---|---|
| TRIAZOLAM | NERVOUS SYSTEM | PSYCHOLEPTICS | HYPNOTICS AND SEDATIVES | HYPNOTICS |
| UBROGEPANT | NERVOUS SYSTEM | ANALGESICS | ANTIMIGRAINE PREPARATIONS | ANALGESICS |
| VALBENAZINE TOSILATE | NERVOUS SYSTEM | OTHER NERVOUS SYSTEM DRUGS | OTHER NERVOUS SYSTEM DRUGS | OTHER |
| VALPROIC ACID | NERVOUS SYSTEM | ANTIEPILEPTICS | ANTIEPILEPTICS | ANTIEPILEPTICS |
| VARENICLINE | NERVOUS SYSTEM | OTHER NERVOUS SYSTEM DRUGS | DRUGS USED IN ADDICTIVE DISORDERS | OTHER |
| VENLAFAXINE | NERVOUS SYSTEM | PSYCHOANALEPTICS | ANTIDEPRESSANTS | ANTIDEPRESSANTS |
| VERAPAMIL | NERVOUS SYSTEM | ANALGESICS | ANTIMIGRAINE PREPARATIONS | ANALGESICS |
| VILAZODONE | NERVOUS SYSTEM | PSYCHOANALEPTICS | ANTIDEPRESSANTS | ANTIDEPRESSANTS |
| VILOXAZINE | NERVOUS SYSTEM | PSYCHOANALEPTICS | ANTIDEPRESSANTS | STIMULANTS |
| VORTIOXETINE | NERVOUS SYSTEM | PSYCHOANALEPTICS | ANTIDEPRESSANTS | ANTIDEPRESSANTS |
| ZALEPLON | NERVOUS SYSTEM | PSYCHOLEPTICS | HYPNOTICS AND SEDATIVES | HYPNOTICS |
| ZIPRASIDONE | NERVOUS SYSTEM | PSYCHOLEPTICS | ANTIPSYCHOTICS | ANTIPSYCHOTICS |
| ZOLMITRIPTAN | NERVOUS SYSTEM | ANALGESICS | ANTIMIGRAINE PREPARATIONS | ANALGESICS |
| ZOLPIDEM | NERVOUS SYSTEM | PSYCHOLEPTICS | HYPNOTICS AND SEDATIVES | HYPNOTICS |
| ZONISAMIDE | NERVOUS SYSTEM | ANTIEPILEPTICS | ANTIEPILEPTICS | ANTIEPILEPTICS |
