## Supplementary material for "Durability of the Benefit of Vagus Nerve Stimulation in Markedly Treatment-Resistant Major Depression: A RECOVER Trial Report": List of Institutional Review Boards

LivaNova
Health innovation that matters

RECOVER
A VNS Therapy Clinical Trial

### INSTITUTIONAL REVIEW BOARDS

**Below is the list of governing IRBs from the sites listed in the acknowledgement section of the manuscript. To date all IRBs in which the study was submitted have been approved. Advarra IRB serves as the central IRB oversight on the RECOVER study.**

| IRB & Address | Study Site | Site Address | IRB Decision |
|---|---|---|---|
| WCG IRB<br>212 Carnegie Center, Suite 301<br>Princeton, NJ 08540 | Emory University | 101 Woodruff Circle NE, Suite 4306<br>Atlanta, GA 30322<br>and<br>2701 North Decatur Rd., Suite 800<br>Decatur, GA 30033 | Approved |
| Advarra<br>6100 Merriweather Dr., Suite 600<br>Columbia, MD 21044 | Psych Atlanta | 1012 Coggins Place<br>Marietta, GA 30060 | Approved |
| Advarra<br>6100 Merriweather Dr., Suite 600<br>Columbia, MD 21044 | Dell Medical School | 1601 Trinity St.<br>Austin, TX 78712 | Approved |
| Advarra<br>6100 Merriweather Dr., Suite 600<br>Columbia, MD 21044 | Icahn School of Medicine at Mount Sinai | One Gustave L. Levy Place<br>New York, NY 10029<br>and<br>1399 Park Avenue, 2nd Floor<br>New York, NY 10029 | Approved |
| Advarra<br>6100 Merriweather Dr., Suite 600<br>Columbia, MD 21044 | Alivation Research, LLC | 8550 Cuthills Circle, Suite 100<br>Lincoln, NE 68526 | Approved |
| TTUHSC El Paso IRB<br>137 Rick Francis Dr.<br>MSBII, Room 5A111A/B<br>El Paso, Texas 79905<br>Mailing: 5001 El Paso Drive MSC Number 31004<br>El Paso, Texas 79905 | Texas Tech University Health Science Center El Paso | 5001 El Paso Drive<br>El Paso, TX 79905 | 1840013 |
| Rush University Medical Center IRB<br>1653 West Congress Parkway<br>Chicago, IL 60612 | Rush University Medical Center<br>Treatment Research Center | 1645 W. Jackson Blvd.<br>Suite 600<br>Chicago, IL 60612 | 1840015 |
| Advarra<br>6100 Merriweather Dr., Suite 600<br>Columbia, MD 21044 | Massachusetts General Hospital | One Bowdoin Square,<br>6th Floor<br>Boston, MA 02114 | Approved |

| IRB & Address | Study Site | Site Address | IRB Decision |
|---|---|---|---|
| University of Pennsylvania IRB<br>3600 Civic Center Blvd., 9th Floor<br>Philadelphia, PA 19104 | University of Pennsylvania Department of Psychiatry | 3700 Hamilton Walk,<br>Suite 303<br>Philadelphia, PA 19104 | Approved |
| Advarra<br>6100 Merriweather Dr., Suite 600<br>Columbia, MD 21044 | dTMS Center, LLC | 11903 Southern Blvd. #104<br>Royal Palm Beach, FL 33411<br>and<br>249 Peruvian Ave. Palm Beach, FL 33480<br>and<br>790 Juno Ocean Walk #404C<br>Juno Beach, FL 33408 | Approved |
| Advarra<br>6100 Merriweather Dr., Suite 600<br>Columbia, MD 21044 | The University of Texas Health Science Center at Houston | 1941 East Road, Suite 2100<br>Houston, TX 77054 | Approved |
| Advarra<br>6100 Merriweather Dr., Suite 600<br>Columbia, MD 21044 | Medical University of South Carolina/ Brain Stimulation Lab Institute of Psychiatry | 67 President Street<br>Charleston, SC 29425 | Approved |
| Advarra<br>6100 Merriweather Dr., Suite 600<br>Columbia, MD 21044 | Sheppard Pratt Health System, Inc. | Clinical Research Programs, A319<br>6501 N. Charles Street<br>Baltimore, MD 21204 | Approved |
| Advarra<br>6100 Merriweather Dr., Suite 600<br>Columbia, MD 21044 | University of Southern California Neurorestoration Center | 1333 San Pablo Street<br>Los Angeles, CA 90033 | Approved |
| Advarra<br>6100 Merriweather Dr., Suite 600<br>Columbia, MD 21044 | Stony Brook Psychiatry | 100 Nicolls Rd.<br>Stony Brook, NY 11794 | Approved |
| Advarra<br>6100 Merriweather Dr., Suite 600<br>Columbia, MD 21044 | PsychCare Consultants Research | 5000 Cedar Plaza Parkway, Suite 220A<br>St. Louis, MO 63128 | Approved |
| Advarra<br>6100 Merriweather Dr., Suite 600<br>Columbia, MD 21044 | University of California, San Diego Medical Center - Hillcrest Hospital | 200 West Arbor Drive<br>San Diego, CA 92103 | Approved |
| Advarra<br>6100 Merriweather Dr., Suite 600<br>Columbia, MD 21044 | University of Minnesota St. Louis Park Clinic | 5775 Wayzata Blvd.,<br>Park Place East Floor 2, | Approved |

| IRB & Address | Study Site | Site Address | IRB Decision |
| --- | --- | --- | --- |
| | | Suite 255<br>St. Louis Park, MN 55416 | |
| Advarra<br>6100 Merriweather Dr., Suite 600<br>Columbia, MD 21044 | ATP Clinical Research, Inc. | 3151 Airway Avenue,<br>Suite T-3<br>Costa Mesa, CA 92626 | Approved |
| Advarra<br>6100 Merriweather Dr., Suite 600<br>Columbia, MD 21044 | Center For Anxiety and Depression | 7525 SE 24th Street,<br>Suite 400<br>Mercer Island, WA 98040 | Approved |
| Advarra<br>6100 Merriweather Dr., Suite 600<br>Columbia, MD 21044 | Augusta University Department of Psychology & Health Behavior | 997 St. Sebastian Way<br>Augusta, GA 30912 | Approved |
| Advarra<br>6100 Merriweather Dr., Suite 600<br>Columbia, MD 21044 | University of Missouri Dept. of Psychiatry, Neuromodulation Clinic | 551 East Southampton Dr.<br>Columbia, MO 65201 | Approved |
| Advarra<br>6100 Merriweather Dr., Suite 600<br>Columbia, MD 21044 | Galiz Research, LLC | 7100 West 20th Ave.,<br>Suite 802<br>Hialeah, FL 33016 | Approved |
| Advarra<br>6100 Merriweather Dr., Suite 600<br>Columbia, MD 21044 | APG Research, LLC | 721 N. Magnolia Ave.<br>Orlando, FL 32803 | Approved |
| SIU Medicine IRB<br>201 East Madison Street<br>P.O. Box 19664<br>Springfield, IL 62794 | Southern Illinois University Medicine | 319 E. Madison St.<br>Springfield, IL 62701 | Approved |
| Advarra<br>6100 Merriweather Dr., Suite 600<br>Columbia, MD 21044 | Hapworth Research, Inc. | 5 East 57th Street, 18th Floor<br>New York, NY 10022 | Approved |
| Advarra<br>6100 Merriweather Dr., Suite 600<br>Columbia, MD 21044 | Neuropsychiatric Associates, Plc | Woodstock Research Center<br>21704 Maxham Meadow Way<br>Woodstock, VT 05091 | Approved |
| Advarra<br>6100 Merriweather Dr., Suite 600<br>Columbia, MD 21044 | Michigan Clinical Research Institute, PC | 3001 Plymouth Road, Suite 107<br>Ann Arbor, MI 48105 | Approved |

| IRB & Address | Study Site | Site Address | IRB Decision |
|---|---|---|---|
| Advarra<br>6100 Merriweather Dr.,<br>Suite 600<br>Columbia, MD 21044 | Seattle Neuropsychiatric Treatment Center | 805 Madison St.,<br>Suite 401<br>Seattle, WA 98104<br>and<br>1450 114th Ave. SE, #110<br>Bellevue, WA 98004 | Approved |
| WCG IRB<br>212 Carnegie Center,<br>Suite 301<br>Princeton, NJ 08540 | Carilion Clinic | 4434 Electric Road<br>Roanoke, VA 24018 | Approved |
| The University of Utah IRB<br>75 South 2000 East<br>Salt Lake City, UT 84112 | The University of Utah | 501 Chipeta Way<br>Salt Lake City, UT 84108 | Approved |
| Advarra<br>6100 Merriweather Dr.,<br>Suite 600<br>Columbia, MD 21044 | AMR - Baber Research, Inc. | 1460 Bond Street,<br>Suite 130<br>Naperville, Illinois 60563 | Approved |
| Advarra<br>6100 Merriweather Dr.,<br>Suite 600<br>Columbia, MD 21044 | Syrentis Clinical Research | 1401 N. Tustin Ave.,<br>Suite 130<br>Santa Ana, CA 92705 | Approved |
| Advarra<br>6100 Merriweather Dr.,<br>Suite 600<br>Columbia, MD 21044 | Washington University | 660 South Euclid Avenue<br>St. Louis, MO 63110 | Approved |
| Advarra<br>6100 Merriweather Dr.,<br>Suite 600<br>Columbia, MD 21044 | University of Wisconsin School of Medicine and Public Health Wisconsin Psychiatric Institute and Clinic Department of Psychiatry | 6001 Research Park Blvd.<br>Madison, WI 53719 | Approved |
| Advarra<br>6100 Merriweather Dr.,<br>Suite 600<br>Columbia, MD 21044 | Precise Research Centers | 3531 Lakeland Drive,<br>Suite 1060<br>Flowood, MS 39232 | Approved |
| WCG IRB<br>212 Carnegie Center,<br>Suite 301<br>Princeton, NJ 08540 | University of Alabama at Birmingham | 1720 7th Avenue South<br>Sparks Center, 10th Floor<br>Birmingham, Alabama 35294 | Approved |
| Advarra<br>6100 Merriweather Dr.,<br>Suite 600<br>Columbia, MD 21044 | Stedman Clinical Trials | 14506 University Point Place<br>Tampa, FL 33613 | Approved |

| IRB & Address | Study Site | Site Address | IRB Decision |
|---|---|---|---|
| Advarra<br>6100 Merriweather Dr., Suite 600<br>Columbia, MD 21044 | Global Medical Institutes, LLC | 340 Montage Mountain Road<br>Moosic, PA 18507 | Approved |
| WCG IRB<br>212 Carnegie Center, Suite 301<br>Princeton, NJ 08540 | The Ohio State University | 1670 Upham Drive<br>Columbus, OH 43210 | Approved |
| Advarra<br>6100 Merriweather Dr., Suite 600<br>Columbia, MD 21044 | Beacon Medical Group Behavioral Health South Bend | 707 N. Michigan St<br>Suite 400<br>South Bend, IN 46601 | Approved |
| Advarra<br>6100 Merriweather Dr., Suite 600<br>Columbia, MD 21044 | Trinity Medical | 5320 Military Road, Suite 106<br>Lewiston, NY 14092 | Approved |
| Advarra<br>6100 Merriweather Dr., Suite 600<br>Columbia, MD 21044 | Mindful Behavioral Health PLLC | 2201 NW Corporate Blvd.<br>Boca Raton, FL 33431 | Approved |
| Advarra<br>6100 Merriweather Dr., Suite 600<br>Columbia, MD 21044 | Neuroscience & TMS Treatment Center | 5300 Maryland Way,<br>Suite 103<br>Brentwood, TN 37027 | Approved |
| Advarra<br>6100 Merriweather Dr., Suite 600<br>Columbia, MD 21044 | Florida Center for TMS | 17 Saint Johns Medical Park Drive<br>Saint Augustine, FL 32086 | Approved |
| Advarra<br>6100 Merriweather Dr., Suite 600<br>Columbia, MD 21044 | SF-Care, Inc | 1330 Lincoln Avenue<br>Suite 308<br>San Rafael, CA 94901 | Approved |
| Advarra<br>6100 Merriweather Dr., Suite 600<br>Columbia, MD 21044 | Signature Research Associates, Inc. | 2820 W. Market Street,<br>Suite 110<br>Fairlawn, OH 44333 | Approved |
| Advarra<br>6100 Merriweather Dr., Suite 600<br>Columbia, MD 21044 | Rakesh Ranjan, M.D. & Associates, Inc.<br>Charak Clinical Research Center | 12395 McCracken Road,<br>Suite E<br>Garfield Heights, OH 44125 | Approved |
| University of Oklahoma IRB<br>865 Research Parkway, Suite 400<br>Oklahoma City, OK 73104 | University of Oklahoma School of Community Medicine | 4444 E. 41st Street<br>Tulsa, OK 74135 | Approved |
| Advarra<br>6100 Merriweather Dr., Suite 600<br>Columbia, MD 21044 | Psychiatric Care and Research Center | 4132 Keaton Crossing Blvd.,<br>Suite 201<br>O'Fallon, MO 63368 | Approved |

| IRB & Address | Study Site | Site Address | IRB Decision |
|---|---|---|---|
| Advarra<br>6100 Merriweather Dr., Suite 600<br>Columbia, MD 21044 | Healthy Perspectives - Innovative Mental Health Services, PLLC | 30 Temple Street, Suite 105<br>Nashua, NH 03060 | Approved |
| Advarra<br>6100 Merriweather Dr., Suite 600<br>Columbia, MD 21044 | Dent Neurologic Institute | 3980 Sheridan Dr., Suites 500, 304, 308, 3980A-Tower A<br>Amherst, NY 14226 | Approved |
| Advarra<br>6100 Merriweather Dr., Suite 600<br>Columbia, MD 21044 | Marshall Clinical Research Center | 1600 Medical Center Drive, Suite 250<br>Huntington, WV 25701 | Approved |
| Advarra<br>6100 Merriweather Dr., Suite 600<br>Columbia, MD 21044 | Florida Behavioral Medicine | 1100 Clearwater Largo Rd. N.<br>Largo, FL 33770 | Approved |
| WCG IRB<br>212 Carnegie Center, Suite 301<br>Princeton, NJ 08540 | University of Alabama at Birmingham Huntsville Regional Medical Center | 301 Governors Dr. SW<br>Huntsville, AL 35801 | Approved |
| Advarra<br>6100 Merriweather Dr., Suite 600<br>Columbia, MD 21044 | Florida Center for TMS | 142 East Gore Street<br>Orlando, FL 32806 | Approved |
| Advarra<br>6100 Merriweather Dr., Suite 600<br>Columbia, MD 21044 | Nova Psychiatry, Inc. | 1836 Woodward Street<br>Orlando, FL 32803 | Approved |
| Advarra<br>6100 Merriweather Dr., Suite 600<br>Columbia, MD 21044 | Kaizen Brain Center | 4180 La Jolla Village Dr., Suite 240<br>La Jolla, CA 92037 | Approved |
| Advarra<br>6100 Merriweather Dr., Suite 600<br>Columbia, MD 21044 | Center for Neuropsychiatry and Brain Stimulation (CNBS) | 351 Wellesley Trade Lane, Suite 201<br>Cary, NC 27519 | Approved |
